# Time trends in social contacts before and during the COVID-19 pandemic: the CONNECT study

**DOI:** 10.1101/2021.10.06.21264632

**Authors:** Mélanie Drolet, Aurélie Godbout, Myrto Mondor, Guillaume Béraud, Léa Drolet-Roy, Philippe Lemieux-Mellouki, Alexandre Bureau, Éric Demers, Marie-Claude Boily, Chantal Sauvageau, Gaston De Serres, Niel Hens, Philippe Beutels, Benoit Dervaux, Marc Brisson

## Abstract

**Background:** Since the beginning of the COVID-19 pandemic, many countries, including Canada, have adopted unprecedented physical distancing measures such as closure of schools and non-essential businesses, and restrictions on gatherings and household visits. We described time trends in social contacts for the pre-pandemic and pandemic periods in Quebec, Canada.

**Methods:** CONNECT is a population-based study of social contacts conducted shortly before (2018/2019) and during the COVID-19 pandemic (April 2020 – February 2021), using the same methodology for both periods. We recruited participants by random-digit-dialing and collected data by self-administered web-based questionnaires. Questionnaires documented socio-demographic characteristics and social contacts for two assigned days. A contact was defined as a two-way conversation at a distance ≤2 meters or as a physical contact, irrespective of masking. We used weighted generalized linear models with a Poisson distribution and robust variance (taking possible overdispersion into account) to compare the mean number of social contacts over time by characteristics.

**Results:** A total of 1291 and 5516 Quebecers completed the study before and during the pandemic, respectively. Contacts significantly decreased from a mean of 8 contacts/day prior to the pandemic to 3 contacts/day during the spring 2020 lockdown. Contacts remained lower than the pre-COVID period thereafter (lowest=3 contacts/day during the Christmas 2020/2021 holidays, highest=5 in September 2020). Contacts at work, during leisure activities/other locations, and at home with visitors showed the greatest decreases since the beginning of the pandemic. All sociodemographic subgroups showed significant decreases of contacts since the beginning of the pandemic. The mixing matrices illustrated the impact of public health measures (e.g. school closure, gathering restrictions) with fewer contacts between children/teenagers and fewer contacts outside of the three main diagonals of contacts between same-age partners/siblings and between children and their parents.

**Conclusion:** Physical distancing measures in Quebec significantly decreased social contacts, which most likely mitigated the spread of COVID-19.

## BACKGROUND

On September 1^st^ 2021, Canada surpassed 1.5 million confirmed cases of COVID-19, and >25% of these cases were from Quebec[1]. While the province of Quebec was the epicenter of the first wave, most Canadian provinces experienced stronger second and third waves in terms of cases and hospitalisations. Since the beginning of the pandemic, Canada has adopted unprecedented physical distancing measures from complete lockdowns to a combination of school and non-essential businesses closures and restrictions on gatherings and household visits, depending on epidemiological indicators and regions[2].

Given that physical distancing measures are a cornerstone of public health COVID-19 mitigation efforts, it is important to examine how social contacts changed over time: 1) to better understand the dynamics of the pandemic, 2) to inform future measures, and 3) to provide crucial data for mathematical modeling. To our knowledge, this is one of the few population-based studies that has compared social contacts documented shortly before and during the COVID-19 pandemic using the same methodology[3].

The main objective of this study is to describe the time trends in social contacts for the COVID-19 pre-pandemic (2018-2019) and pandemic periods (April 2020-February 2021) in Quebec, Canada using a social contact survey and a representative sample of the population. Specific objectives are to describe the time trends in the number of social contacts, overall and by location (home, work, school, public transport, leisure, other) and by key socio-demographic characteristics.

## METHODS

### Study design

CONNECT (CONtact and Network Estimation to Control Transmission) is a population-based survey of epidemiologically relevant social contacts and mixing patterns conducted in the province of Quebec, Canada. The first phase of CONNECT was conducted in 2018-2019 (February 2018 to March 2019), one year before the COVID-19 pandemic. Four additional phases of CONNECT were undertaken to document changes in social contacts during the COVID-19 pandemic period (CONNECT2: April 21^st^-May 25^th^ 2020 and CONNECT3,4,5: July 3^rd^ 2020-February 26^th^ 2021) (Additional file 1: Table S1). All CONNECT phases were conducted with the same methodology.

### Recruitment of participants

The target population of CONNECT consisted of all non-institutionalized Quebecers without any age restriction (e.g., elderly living in retirement homes who generally have personal phone lines were eligible but those living in long-term care homes (nursing homes, Quebec CHSLD) were not eligible). We used random digit dialling to recruit participants. The randomly generated landline and mobile phone number sample was provided by ASDE, a Canadian firm specialized in survey sampling [4]. After having explained the study, verified eligibility of the household and documented the age and sex of all household members, we randomly selected one person per household to participate in the study, using a probability sample stratified by age. This recruitment procedure was sequentially repeated for every new phase of CONNECT (i.e, new participants were recruited for every CONNECT phase).

### Data collection

We collected data using a self-administered web-based questionnaire. A secured individualized web link to the questionnaire and information about the study were sent by email to each selected participant who consented to participate in the study. Parents of children aged less than 12 years were asked to complete the questionnaire on behalf of their child, whereas teenagers aged 12-17 years completed their own questionnaire, after parental consent.

The same questionnaire was used for all CONNECT phases. The first section of the questionnaire documented key socio-demographic characteristics. The second section was a social contact diary, based on instruments previously used in Polymod and other similar studies[5-7] (an example of the diary is provided in the Additional file 1: Figure S1). Briefly, participants were assigned two random days of the week (one week day and one weekend day) to record every different person they had contact with between 5 am and 5 am the following morning. A contact was defined as either physical (handshake, hug, kiss) or nonphysical (two-way conversation in the physical presence of the person, at a distance equal or less than 2 meters, irrespective of masking). Participants provided the characteristics of the contact persons (age, sex, ethnicity, and relationship to themselves (e.g., household member, friend, colleague)) as well as characteristics of the contacts with this person: location where the contact(s) occurred (home, work, daycare/school, public transport, leisure, other location), duration, usual frequency of contact with that person, and whether the contact was physical or not. Participants reporting more than 20 professional contacts per day were asked not to report all their professional contacts in the diary. Instead they were asked general questions about these professional contacts: age groups of the majority of contact persons, average durations of contacts and whether physical contacts were generally involved or not. Additional questions about teleworking were included from CONNECT2 onwards.

All CONNECT phases were approved by the ethics committee of the CHU de Québec research center (project 2016-2172) and we commissioned the market company Advanis for recruitment and data collection. All participants gave their consent to participate in the study during the recruitment phone call. Informed consent was taken from a parent and/or legal guardian for study participation in the case of minors.

### Analyses

We weighted the participants of the CONNECT 1-5 surveys by age, sex, region (Greater Montreal and other Quebec regions), and household composition (households without 0-17-year-olds, households with 0-5-year-olds, if not with 6-14-year-olds, if not with 15-17-year-olds), using the Quebec data of the 2016 Canadian census (Additional file 1: Table S2) and we verified that they were representative of the Quebec population for key socio-demographic characteristics. To obtain daily number of social contacts on a weekly basis, we weighted the number of daily contacts reported during the week (5/7) and the weekend (2/7). We classified the type of employment of workers using the 2016 National occupation classification (NOC)[8].

We estimated the number of social contacts per person and per day, for all locations combined and for 6 different locations: home, work, school, public transportation, leisure, and other locations. To do so, several steps were necessary. First, for a contact person met in more than a single location during a single day, the location of the contact was assigned in the following hierarchical order, according to risk of transmission: home, work, school, public transport, leisure and other locations [9]. For example, if a parent reported contacts with his child at home, in public transportation and in a leisure activity, we only considered the home contact to avoid counting contacts with the same person multiple times. Second, for workers reporting more than 20 professional contacts per working day, we added their reported number of professional contacts to the work location for their working day(s). Similar to other studies which allowed a maximal number of contacts per day [5, 6, 10], we truncated professional contacts at a maximum of 40 per day to eliminate extreme values and contacts at low risk of transmission of infectious diseases. Third, we identified all workers in schools through their NOC code and job descriptions and attributed their professional contacts to the school location. We did so to describe social contacts in schools, not only between students, but also between students and their teachers, educators, and other school’s workers. Unless specified, we estimated the mean number of contacts in the different locations using a population-based denominator. With this method, all individuals were considered in the denominator of each location had they reported contacts or not for that location. The sum of contacts in the different locations gives the total number of contacts.

Using data available from CONNECT1-5, we determined different periods to reflect the Quebec COVID-19 epidemiology, their related physical distancing measures, and expected seasonality in social contacts (Additional file 1: Figure S2). We used data collected from February 1^st^ 2018 to March 17^th^ 2019 as our pre-COVID period. We used data collected from April 21^st^ to May 25^th^ 2020 to represent the first wave, data collected from July 3^rd^ to August 31^st^ 2020 to represent the summer, and data collected from September 1^st^ 2020 to February 26^th^ 2021 to represent the second wave. We further stratified the second wave to represent periods of expected seasonality in social contacts: September with the return to school and at work, fall with gathering restrictions, the Christmas holidays with school and work vacations and closure of non-essential business, January and February 2021 with the gradual return to work and school after Christmas vacations and school/non-essential business closures, and the introduction of a curfew. We used a Canadian stringency index, adapted from the Oxford COVID-19 Government Response Tracker (OxCGRT) [11], to quantify the intensity of public health measures in Quebec over time [12]. This index is obtained by averaging the intensity score of 12 policy indicators (e.g., school closures, workplace closures, gathering restrictions, stay-at-home requirements, etc.) and higher values indicate stricter measures. We estimated the mean stringency index for each of the 8 periods described previously by averaging the daily values of the index.

We compared the mean number of social contacts over time (total or by location) using weighted generalized linear models with a Poisson distribution and an identity link. Generalized estimating equations with robust variance [13] were used to account for the correlation between the two days of diary data collection and overdispersion. A categorical period effect was included in the model and is presented as the absolute difference in the mean number of contacts compared to the previous period. We also compared the mean number of social contacts according to different key socio-demographic characteristics using the same model with a period-by-covariate interaction and adjusting for age (in 8 categories). In this model, period and characteristic effects were tested using contrasts: each period was compared to the previous period within each level of the covariate, and the global effect of the characteristic was tested within each period.. We also examined the association between the mean number of social contacts and the stringency index (in 5 categories), irrespective of periods, using a model similar to the one comparing periods.

Finally, we estimated mixing matrices. The entries of the mixing matrix represent the mean number of social contacts per person per day according to the age of the respondent (column) and the age of his contacts (row). Mixing matrices were estimated separately for the 8 periods described previously and for 3 categories of contact locations: all locations, home (contacts with household members and visitors), any location outside home. The matrices were obtained by maximizing a constrained log likelihood of the number of reported contacts per day among CONNECT participants weighted by age, sex, household composition and region. The number of contacts was assumed to follow a negative binomial distribution. The likelihood constraint ensured that the total number of contacts between individuals of age i and age j is the same whether it is estimated from entry (i,j) or entry (j,i) of the total mixing matrix including contacts in all locations (i.e., reciprocity of the mixing matrix).

All statistical analyses were performed with SAS version 9.4. Maximization of the log likelihood for the mixing matrices was performed using a nonlinear programming algorithm (nlminb2 function from the ROI package in R).

## RESULTS

### Participants

A total of 1291, 546, and 4970 Quebecers completed the social contact questionnaires during the pre-COVID period (CONNECT1), the first wave (CONNECT2), and summer 2020 and second wave (CONNECT3-5), respectively. Participation rates (number of questionnaires completed among consenting participant) were 30%, 38%, and 34% for CONNECT 1,2, and 3-5 respectively (Additional file 1: Figure S3). These participants were generally representative of the Quebec general population, and they were comparable across the different phases of CONNECT (Table 1).

**Table 1.** Key socio-demographic characteristics of CONNECT participants and the Quebec general population.

|  | 2016<br>Census | Pre-COVID |  | 1<sup>st</sup> wave |  | Summer 2020<br>and 2<sup>nd</sup> wave |  |
| --- | --- | --- | --- | --- | --- | --- | --- |
|  | % | N | % weighted | N | % weighted | N | % weighted |
| **Total** |  | **1291** |  | **546** |  | **4970** |  |
| **Age** |  |  |  |  |  |  |  |
| 0-5 yrs old | 6.2 | 222 | 6.6 | 40 | 8.4 | 298 | 7.0 |
| 6-11 yrs old | 6.6 | 163 | 8.1 | 31 | 5.7 | 225 | 7.5 |
| 12-17 yrs old | 5.9 | 91 | 6.8 | 60 | 8.0 | 506 | 7.3 |
| 18-25 yrs old | 9.3 | 98 | 8.5 | 53 | 9.1 | 506 | 9.3 |
| 26-45 yrs old | 26.4 | 204 | 27.9 | 181 | 26.3 | 1304 | 26.8 |
| 46-65 yrs old | 27.6 | 303 | 25.8 | 131 | 25.8 | 1539 | 26.0 |
| 66-75 yrs old | 10.6 | 162 | 9.7 | 45 | 9.8 | 514 | 9.7 |
| >75 yrs old | 7.4 | 48 | 6.4 | 5 | 6.8 | 78 | 6.5 |
| **Sex** |  |  |  |  |  |  |  |
| Male | 49.6 | 609 | 49.9 | 238 | 49.2 | 2515 | 50.0 |
| Female | 50.4 | 682 | 50.1 | 308 | 50.8 | 2455 | 50.0 |
| **Region** |  |  |  |  |  |  |  |
| Rural | 18.8 | 239 | 15.5 | 66 | 12.1 | 836 | 16.2 |
| Urban | 81.2 | 1049 | 84.5 | 480 | 87.9 | 4134 | 83.8 |
| **Region** |  |  |  |  |  |  |  |
| Greater Montreal<sup>e</sup> | 61.0 | 635 | 61.0 | 371 | 61.0 | 2815 | 60.9 |
| Other Quebec regions | 39.0 | 642 | 39.0 | 175 | 39.0 | 2152 | 39.1 |
| **Household size** |  |  |  |  |  |  |  |
| 1 | 33.3 | 239 | 23.2 | 125 | 23.3 | 968 | 19.6 |
| 2 | 34.8 | 408 | 34.1 | 188 | 36.2 | 2049 | 39.6 |
| 3 | 13.9 | 198 | 13.4 | 78 | 13.1 | 738 | 15.5 |
| 4 | 12.1 | 268 | 17.1 | 108 | 18.9 | 824 | 17.8 |
| 5+ | 6.0 | 178 | 12.3 | 47 | 8.5 | 391 | 7.6 |
| **Household composition** |  |  |  |  |  |  |  |
| Without 0-17-year-olds | 61.0 | 734 | 65.2 | 346 | 65.2 | 3345 | 64.8 |
| With 0-5-year-olds | 17.1 | 256 | 16.2 | 84 | 16.4 | 743 | 16.6 |
| If not, with 6-14-year-olds | 14.0 | 255 | 16.1 | 90 | 15.8 | 734 | 16.0 |
| If not, with 15-17-year-olds | 2.6 | 46 | 2.5 | 26 | 2.5 | 148 | 2.6 |
| 0-17 without information | 5.3 | -- | -- | -- | -- | -- | -- |
| **Level of education (among 25-64 yrs)** |  |  |  |  |  |  |  |
| No diploma, no degree | 13.3 | 38 | 7.7 | 18 | 6.8 | 112 | 3.7 |
| Secondary (high) school | 18.5 | 63 | 12.1 | 34 | 12.5 | 293 | 10.1 |
| College, cegep, other non-university certificate/diploma | 38.8 | 184 | 37.6 | 99 | 32.3 | 916 | 31.9 |
| University | 29.3 | 210 | 42.7 | 164 | 48.4 | 1483 | 54.2 |
| **Employment rate** |  |  |  |  |  |  |  |
| among 15-19 yrs old | 47.3 | 10 | 22.9 | 8 | 22.2 | 98 | 31.7 |
| among 20-24 yrs old | 72.1 | 37 | 42.5 | 20 | 55.4 | 160 | 51.1 |
| among 25-44 yrs old | 85.2 | 179 | 81.8 | 147 | 83.9 | 1183 | 89.6 |
| among 45-64 yrs old | 71.4 | 167 | 59.3 | 99 | 71.0 | 1074 | 72.8 |
| among ≥ 65 yrs old | 10.3 | 22 | 8.6 | 10 | 13.7 | 159 | 19.6 |
|  | % | N | % weighted | N | % weighted | N | % weighted |
| **Participation rate in education** |  |  |  |  |  |  |  |
| among 18-24 yrs old | 55.0 | 51 | 67.9 | 33 | 68.9 | 319 | 70.3 |
| among 25-29 yrs old | 14.0 | 12 | 19.5 | 7 | 16.5 | 66 | 18.2 |
| among 30-34 yrs old | 8.0 | 7 | 15.0 | 5 | 8.5 | 40 | 9.8 |
| **Ethnicity** |  |  |  |  |  |  |  |
| Caucasian | 87.0 | 1124 | 88.5 | 460 | 87.6 | 4449 | 90.3 |
| Other | 13.0 | 156 | 11.5 | 77 | 12.4 | 455 | 9.7 |
| Missing |  | 11 | -- | 9 | -- | 66 | -- |
| **Country of origin** |  |  |  |  |  |  |  |
| Canadian-born | 85.0 | 1208 | 91.5 | 472 | 88.9 | 4498 | 89.8 |
| Foreign-born | 15.0 | 83 | 8.5 | 72 | 11.1 | 466 | 10.2 |
| Missing |  | 0 | -- | 2 | -- | 6 | -- |
| **Mother tongue** |  |  |  |  |  |  |  |
| English | 8.0 | 77 | 7.0 | 43 | 6.8 | 351 | 7.4 |
| French | 79.0 | 1122 | 88.1 | 440 | 84.1 | 4288 | 86.3 |
| Other | 13.0 | 54 | 4.9 | 56 | 9.2 | 291 | 6.3 |
| Missing |  | 38 | -- | 7 | -- | 40 | -- |
| **Type of occupation (workers)\*** |  |  |  |  |  |  |  |
| 0. Management | 9.8 | 52 | 13.3 | 35 | 13.5 | 346 | 12.5 |
| 1. Business, finance, administration | 15.9 | 58 | 13.2 | 56 | 17.4 | 510 | 19.0 |
| 2. Natural & applied sciences | 6.7 | 31 | 8.2 | 33 | 11.6 | 350 | 13.6 |
| 3. Health | 7.0 | 48 | 10.8 | 32 | 9.6 | 224 | 8.3 |
| 4. Education, law & social,<br>community & gov. service | 11.8 | 89 | 20.3 | 48 | 15.4 | 446 | 16.2 |
| 5. Art, culture, recreation & sport | 3.2 | 15 | 3.4 | 12 | 4.1 | 115 | 4.8 |
| 6. Sales & services | 23.2 | 67 | 15.1 | 41 | 12.2 | 382 | 13.7 |
| 7. Trades, transport & equipment<br>operators | 13.5 | 43 | 12.6 | 26 | 11.0 | 240 | 8.6 |
| 8. Natural resources, agriculture &<br>related production | 1.6 | 3 | 1.0 | 3 | 1.4 | 21 | 0.9 |
| 9. Manufacturing & utilities | 4.9 | 6 | 2.0 | 11 | 3.8 | 64 | 2.4 |
| Unknown | 2.4 | 3 | -- | 2 | -- | 48 | -- |

<sup>€</sup> Greater Montreal: Regions of Montréal, Laval, Montérégie, Lanaudière, Laurentides.
\* 2016 National occupation classification.
**Pre-COVID:** February 1<sup>st</sup> 2018 to March 17<sup>th</sup> 2019; **1<sup>st</sup> wave:** April 21<sup>st</sup> to May 25<sup>th</sup> 2020; **Summer 2020 and 2<sup>nd</sup> wave:** July 3<sup>rd</sup> 2020 to February 26<sup>th</sup> 2021.

### Time trends in the number of social contacts

During the pre-pandemic period, the mean number of social contacts per person per day was 7.8 (95% confidence interval (CI):7.2-8.5) (Figure 1 and Additional file 1: Table S3). This number decreased significantly by 60% during the spring 2020 lockdown to 3.1 (95% CI:2.6-3.5). It then increased gradually during summer 2020 and peaked at 5.0 (95% CI:4.3-5.8) contacts/day in September 2020; this peak coincided with the return to school and work. The mean number of contacts decreased significantly again during fall 2020 to 4.1 (95% CI:3.7-4.5) when physical distancing measures were intensified in Quebec to control the second wave. The mean number of social contacts also decreased significantly during the Christmas holidays at 2.9 (95% CI:2.7-3.1) because of school and work vacations and closure of non-essential businesses. There was a trend towards increasing numbers of contacts in January (3.5, 95% CI (3.0-3.9)) with the gradual return to school and in February 2021 (4.0, 95% CI (3.3-4.6)) with the re-opening of non-essential businesses. These time trends in social contacts closely followed the intensity of public health measures as quantified by the stringency index (Figure 1). The mean number of contacts was also significantly associated with the stringency index, irrespective of periods (Additional file 1: Table S4 and Figure S4).

**Figure 1.**
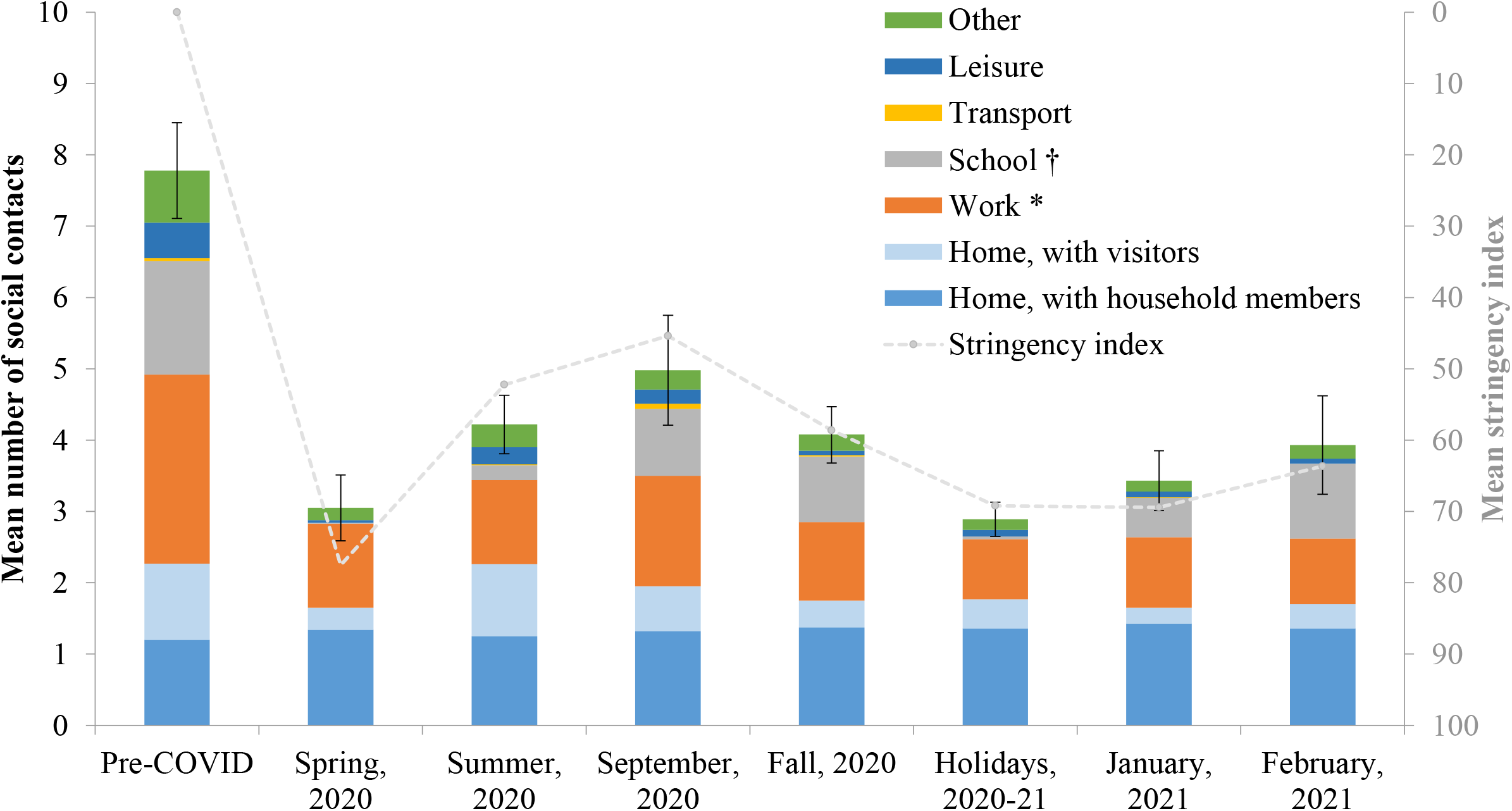
Time trends in the number of social contacts in the province of Quebec. **Pre-COVID:** February 1^st^ 2018 to March 17^th^ 2019**; Spring 2020**: April 21^st^ to May 25^th^ 2020; **Summer 2020**: July 3^rd^ to August 31^st^ 2020; **Fall 2020**: October 1^st^ to December 16^th^ 2020; **Holidays 2020-2021**: December 17^th^ 2020 to January 8^th^ 2021. **1**^**st**^ **wave:** Spring 2020; **2**^**nd**^ **wave**: September 2020 to February 2021. * Contacts at work were truncated to a maximum of 40 contacts per day. ^†^ Contacts for workers in schools were included in the school location. Stringency index: Higher values indicate stricter measures; the mean stringency index for each of the 8 periods was obtained by averaging the daily values of the index. Error bars represent the 95% confidence interval of the total number of social contacts

During the pre-pandemic period, the great majority of contacts occurred at home (2.3 contacts: 1.2 with household members and 1.1 with visitors), at work (2.7 contacts) and at school (1.6 contacts) (Figure 1 and Additional file 1: Table S3). The mean number of contacts at home with household members remained relatively constant over time (1.2 to 1.4 contacts), whereas the number of contacts at home with visitors varied significantly through the study period with lower numbers observed during the spring 2020 lockdown, in January and February 2021 (0.2-0.3 contacts). Compared to the pre-pandemic period, contacts at work and school decreased significantly during the spring lockdown (1.2 and 0.0, respectively), summer (1.2 and 0.2) and the holidays (0.8 and 0.0) and peaked in September 2020 (1.5 and 0.9) with the return at work and school. Contacts in the other locations (transport, leisure and other locations) represented a small proportion of overall contacts during the pre-pandemic period (1.3 contacts). They also decreased significantly since the beginning of the pandemic and stayed low through the study period.

### Time trends in the number of social contacts by age

The location of social contacts varied substantially by age (Figure 2, Additional file 1: Table S5). Contacts in households represented an important part of contacts for all age groups. Contacts in other locations were highly dependent on age. The main locations of contacts away from home for individuals aged 0-17, 18-65, and >65 years were, respectively, school, work, and other locations.

**Figure 2.**
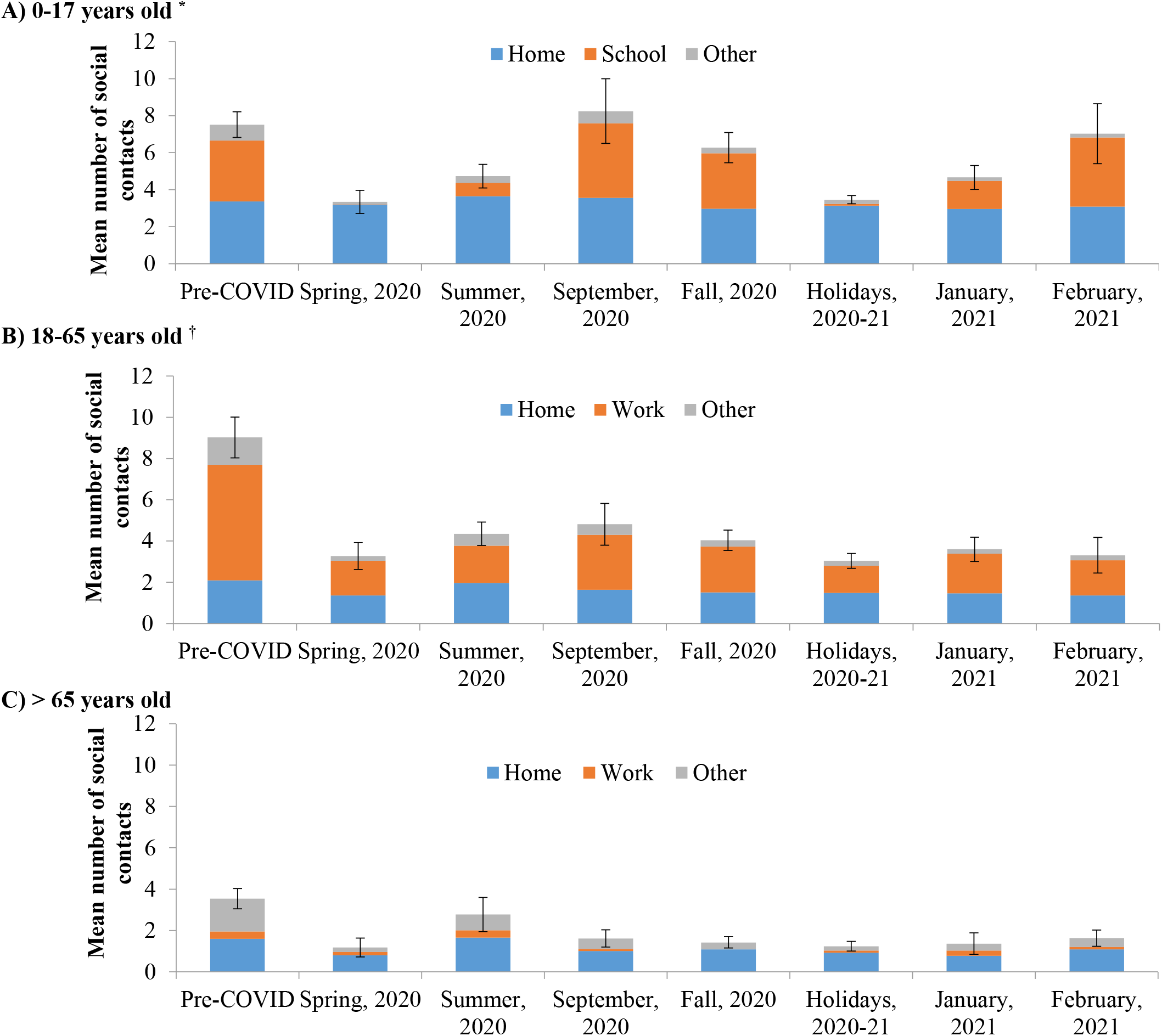
Time trends in the number of social contacts in the province of Quebec, according to age of participants and location of contacts. A) 0-17 years old * B) 18-65 years old ^†^ C) C) > 65 years old **Pre-COVID:** February 1^st^ 2018 to March 17^th^ 2019**; Spring 2020**: April 21^st^ to May 25^th^ 2020; **Summer 2020**: July 3^rd^ to August 31^st^ 2020; **Fall 2020**: October 1^st^ to December 16^th^ 2020; **Holidays 2020-2021**: December 17^th^ 2020 to January 8^th^ 2021. **1**^**st**^ **wave:** Spring 2020; **2**^**nd**^ **wave**: September 2020 to February 2021. * Only the main locations of contacts were included in this figure for 0-17-year-olds (contacts at work were excluded) ^†^ For adults: contacts at work are truncated to a maximum of 40 contacts per day and they include contacts in schools for school workers and students. Contacts in other locations include transports, leisure and other locations. Error bars represent the 95% confidence interval of the total number of social contacts

During the pre-pandemic period, the mean number of social contacts at school/daycare for youth aged 0-17 years was 3.3 contacts (Figure 2A). These contacts significantly decreased to nearly 0 during the spring 2020 lockdown and the Christmas holidays. They reached the pre-pandemic level with the return to school in September (4.0 contacts), during fall 2020 (3.0) and in February 2021 (3.7). Except for post-secondary, similar time trends in contacts at school/daycare were observed by education level (daycare, elementary, high school) (Additional file 1: Table S6).

During the pre-pandemic period, the mean number of contacts at work for adults aged 18-65 years was 5.6 (Figure 2B). These contacts significantly decreased to 1.7 during the spring 2020 lockdown and thereafter remained significantly lower than the pre-pandemic period (from 1.3 during the Christmas holidays to 2.7 in September). The number of contacts at work varied by the type of occupation and the proportion of workers reporting teleworking, and therefore having no contact at work (Additional file 1: Tables S7,S8). During the pre-pandemic period, the greatest number of contacts at work were reported by workers in the domains of Sales & services (10.8), Management (10.2), and Health (10.1). Contacts at work decreased during the spring 2020 lockdown for the majority of domains and remained lower than the pre-pandemic period thereafter. Except for workers in the domains of Health and Sales & Services, the majority of workers in the other domains (> 50%) reported teleworking since the beginning of the pandemic.

During the pre-pandemic period, the mean number of social contacts in other locations for adults older than 65 years was 1.6 (Figure 2C). These contacts decreased significantly at the beginning of the pandemic and remained low through the study period (between 0.2 and 0.8). Therefore, adults older than 65 years had virtually no contact outside their house during this period.

### Time trends in the number of social contacts by key socio-demographic characteristics

During the pre-pandemic period, the mean number of social contacts was significantly higher among individuals living in households with ≥3 individuals (vs households with 1-2 individuals), in households with 0-17-year-olds (vs household without 0-17-year-olds), among native French or English speakers (vs other mother tongues), and among individuals with a university degree (vs no degree) (Figure 3, Additional file 1: Table S5). During the first wave, social contacts significantly decreased for most socio-demographic characteristics. The mean number of social contacts slightly increased after the first wave for all socio-demographic characteristics, although it remained lower than the pre-pandemic period through the study period. During the second wave, the only significant differences between socio-demographic characteristics were a higher number of contacts in households with more individuals and/or households with 0-17-year-olds, mainly explained by the greater number of contacts with household members. Of note, individuals with a university degree had the greatest decrease of their social contacts during the first wave (from 10.7 to 2.5, p<0.0001) and their contacts remained relatively low through the study period (2.5 to 4.4).

**Figure 3.**
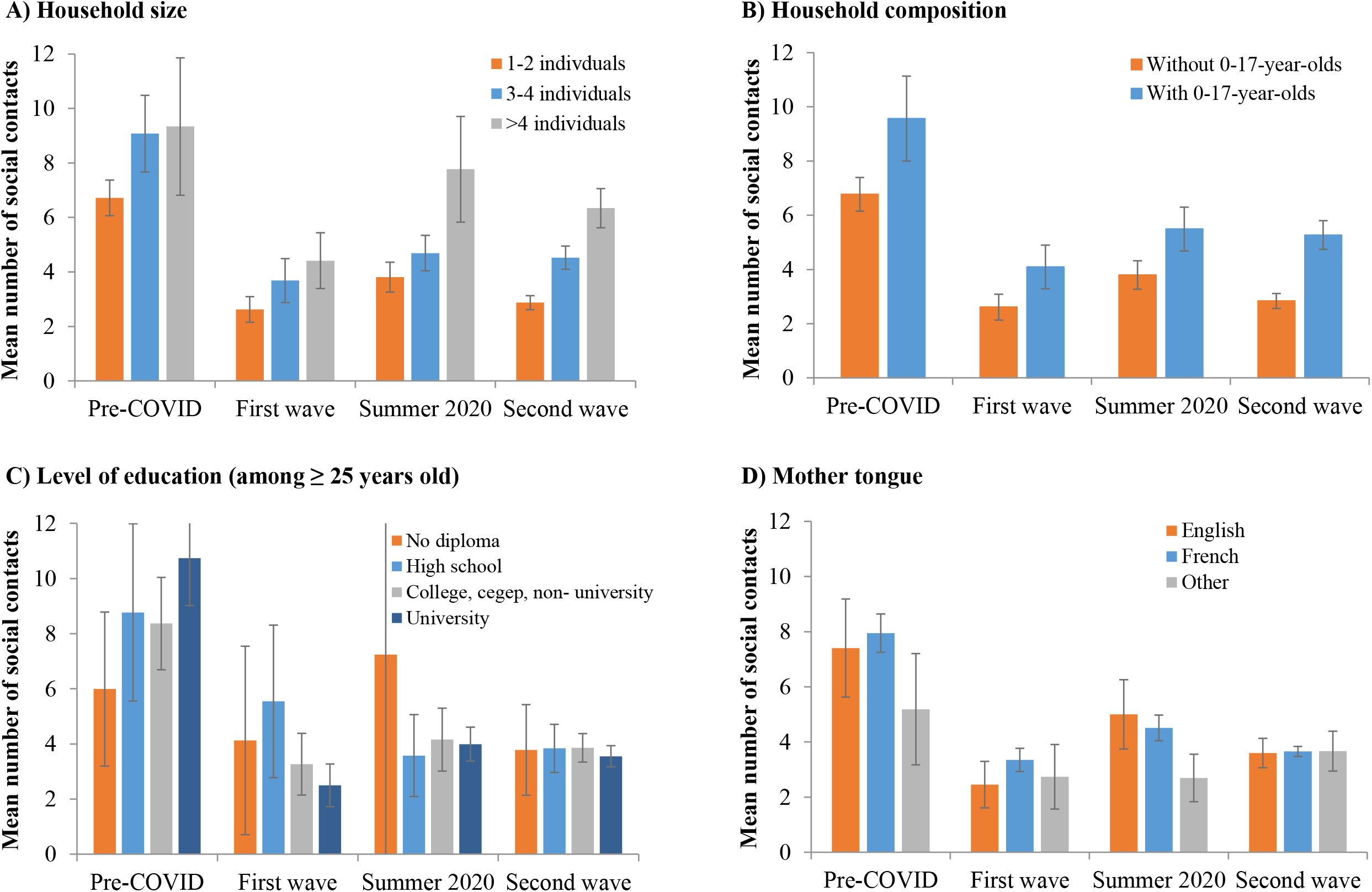
Time trends in the total number of social contacts in the province of Quebec, according to key socio-demographic characteristics (adjusted for age). A) Household size B) Household composition C) Level of education (among ≥ 25 years old) D) Mother tongue **Pre-COVID:** February 1^st^ 2018 to March 17^th^ 2019; **1**^**st**^ **wave:** April 21^st^ to May 25^th^ 2020; **Summer 2020:** July 3^rd^ to August 31^st^ 2020; **2**^**nd**^ **wave:** Sept 1^st^ 2020 to February 26^th^ 2021. Error bars represent the 95% confidence interval of the total number of social contacts

### Time trends in social contact matrices

During the pre-pandemic period, the mixing matrices indicated a high assortativity of contacts by age (as illustrated by the central diagonal), and mixing between children and adults, mainly at home (as illustrated by the 2 secondary diagonals) (Figure 4). These general mixing patterns with 3 diagonals remained apparent during the different pandemic periods, even though the number of contacts was substantially reduced. Interestingly, the matrices of contacts in any location outside home clearly illustrate the impact of school closures or holidays (Spring 2020, Summer 2020, Holidays 2020-21) with fewer contacts between children/teenagers. The matrices of contacts at home (with household members and visitors) also illustrate the impact of restrictions on private gatherings (Spring 2020, Fall 2020 to February 2021) with fewer contacts outside the 3 main diagonals and contacts limited to household members (i.e., same-age partners/siblings and children/parents).

**Figure 4.**
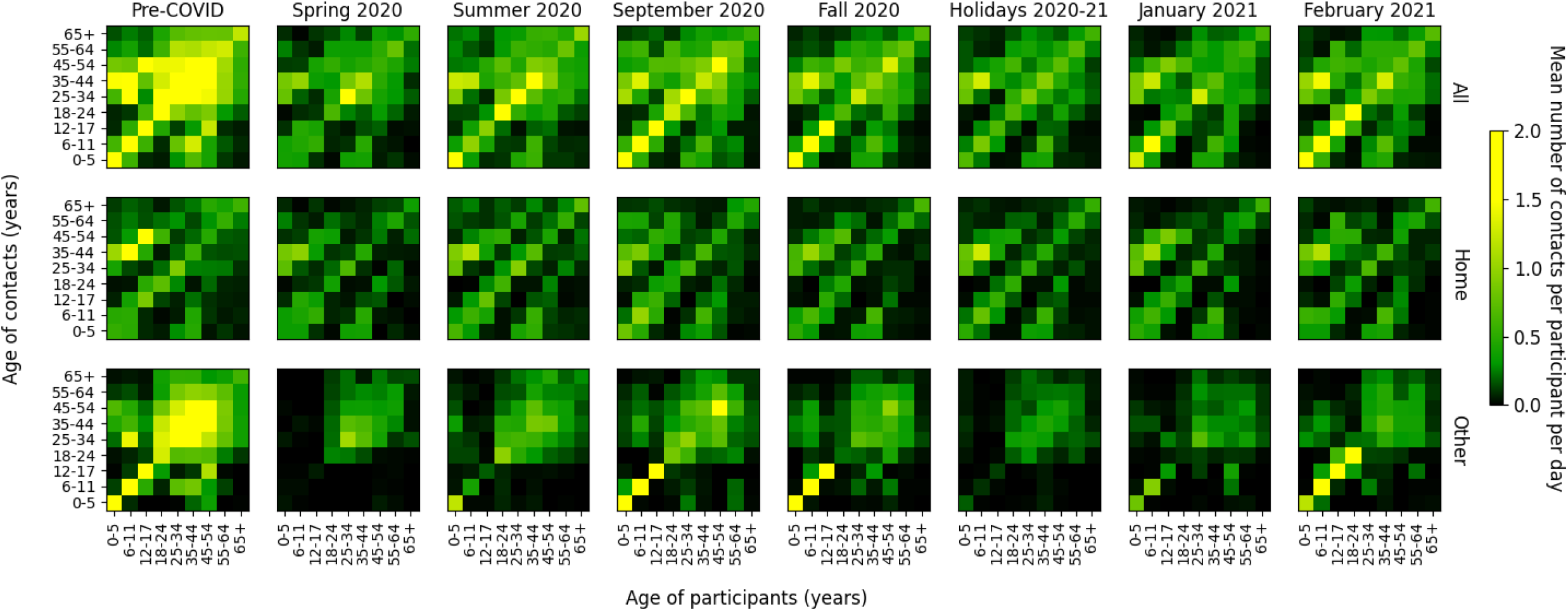
Time trends in social contacts matrices. **Pre-COVID:** February 1^st^ 2018 to March 17^th^ 2019**; Spring 2020**: April 21^st^ to May 25^th^ 2020; **Summer 2020**: July 3^rd^ to August 31^st^ 2020; **Fall 2020**: October 1^st^ to December 16^th^ 2020; **Holidays 2020-2021**: December 17^th^ 2020 to January 8^th^ 2021. **1**^**st**^ **wave:** Spring 2020; **2**^**nd**^ **wave**: September 2020 to February 2021. The matrices of contacts at home include contacts with household members and visitors. The matrices of contacts in other locations include contacts at work, school, transport, leisure, and other locations.

## DISCUSSION

Public health measures to control the COVID-19 spread in Quebec had a significant impact on the number of social contacts. Contacts decreased from a mean of 8 contacts per day prior to the pandemic to 3 contacts per day during the spring 2020 lockdown, a 60% decline. Contacts then increased gradually during the 2020 summer to peak at 5 contacts per day in September with the return to school and at work (36% decline vs pre-COVID). Contacts decreased thereafter during the fall 2020 and winter 2021 to about 4 contacts per day as the physical distancing measures were intensified in Quebec to control the second wave of COVID-19 (47% decline vs pre-COVID). Contacts at work, at school, in leisure activities, and at home with visitors showed the greatest changes through the study period. Before the pandemic, adults aged 18-65 years, individual with a university degree, those living in households with 3 or more individuals and/or in households with 0-17-year-olds, and native French or English speakers reported the greatest number of social contacts. Contacts decreased significantly among all socio-demographic subgroups during the spring 2020 lockdown and remained lower than the pre-pandemic period through the study period.

Our results indicating a 60% reduction of social contacts during the spring 2020 lockdown in Quebec (from 7.8 to 3.1) are generally consistent with the results from similar studies. The CoMix survey, an ongoing empirical study of social contacts conducted in several European countries[14-16], estimated a 70-80% reduction in the number of social contacts during the spring 2020 lockdown compared to similar studies conducted in 2006 (POLYMOD) and 2010 [5, 17]. For example, contacts decreased from 10.8 in 2006[5] to 2.8 contacts per day during the lockdown in United Kingdom[14]. A Canadian study also estimated a 56-80% reduction in social contacts in May, July, September and December 2020[18] compared to POLYMOD data collected in United Kingdom in 2006[5]. However, it is difficult to determine, from these studies, which part of the decrease is related to socio-demographic changes between 2006/2010 and 2020 and to the COVID-19 pandemic. Furthermore, the authors of the Canadian study recognized that social contacts collected in the United Kingdom in 2006 may not be representative of Canadian contacts before the pandemic[18]. Other studies from different countries (e.g. Belgium, France, Germany, Italy, Netherlands, Spain, United Kingdom, United States, Luxembourg, China) have also estimated a mean of around 3 contacts per day during the spring 2020 lockdown period [3, 15, 19-22] and similar increasing trends in social contacts after the first lockdown when physical distancing measures were relaxed[3, 15, 22, 23]. Our results are also consistent with Google phone mobility data for Quebec showing substantial decreases in visits of about 80% in retail & recreation, work, and transit transportation stations during the spring 2020 lockdown compared to January 2020. Mobility increased thereafter but remained lower than the pre-COVID levels for these locations (mean decreases of 20%, 25% and 45% for visits in retail & recreation, work, and transit transportation stations, respectively, from September to Mid-December 2020)[24].

To our knowledge this is one of the few population-based studies of social contacts worldwide to compare social contacts during the pandemic to those documented shortly before the pandemic using the same methodology. Only one other study conducted in the Netherland included social contacts documented shortly before the pandemic (in 2016-2017) and during the pandemic using the same methodology[3]. However, CONNECT has some limitations. Firstly, previous data suggested that social contacts measured with survey methodology could underestimate the number of social contacts compared with a sensors methodology, particularly for contacts of short duration[25, 26]. More specifically, parents participating in CONNECT reported difficulties in reporting contacts at school on behalf of their child. Secondly, although CONNECT is population-based with a random recruitment of the general population, volunteer participants may differ from those refusing to participate in the study and may be those adhering the most to the public health measures. However, we have collected a wealth of information regarding the participant’s characteristics and we are confident that the recruitment process was successful in providing a sample of participants generally representative of the Quebec general population (in terms of region, participation rate to education and employment, race, country of origin and mother tongue), and samples are comparable across the different phases of the study. Thirdly, given that public health measures undertaken aimed at limiting social contacts, social desirability may have contributed to an underestimation of contacts. Some participants may not have reported all their contacts, particularly contacts forbidden by public health measures. These three main limitations would likely bias our results towards an underestimation of social contacts. Nonetheless, changes in social contacts measured in our study closely followed the epidemiology and physical distancing measures in Quebec (Figure 1, Additional file 1: Figures S2 and S4). For example, the beginning of the second wave coincided with an increasing number of social contacts related to school and work return in September. The number of cases stabilisation/decrease of the second wave coincided with a decreasing number of contacts related to the intensification of public health measures in January and February 2021 (Figure 1).

Our results have important implications for COVID-19 control and policy decisions in Quebec and elsewhere. First, continuous monitoring of social contacts represents a measure of the effectiveness of public health measures aiming at reducing social contacts to contain and prevent COVID-19 transmission. Our results suggest that Quebecers have been generally adherent to public health measures since the beginning of the pandemic. For example, restriction of household contacts with visitors was an important public health measure during the spring lockdown and since October 2020 in Quebec. This is clearly reflected by the small number of household contacts with visitors during the spring and fall 2020 and by changes in household mixing matrices with fewer contacts outside of the 3 main diagonals of contacts between same-age partners/siblings and between children and their parents. Second, data on age- and location-specific changes in social contacts and mixing matrices are proxies for contact events that can lead to transmission when made between susceptible and infectious individuals and are an essential input for transmission-dynamic mathematical models considering different types of contacts. Our social contacts data and mixing matrices have been integrated and were regularly updated into our COVID-19 mathematical model for projections of the potential evolution of the pandemic in Quebec to help inform policy decisions [27]. Finally, social contact data can generate hypothesis to improve our understanding of the COVID-19 transmission dynamics. For example, an important increase in the number of cases while the number of contacts remains relatively stable could suggest that the virus became more transmissible per contact. Hypothesis such as the introduction of a new variant more transmissible in a region, new transmission modes, or a higher transmissibility of the virus for specific meteorological conditions could then be explored.

In conclusion, physical distancing measures in Quebec were effective at significantly decreasing social contacts, which most likely helped prevent COVID-19 spread and generalized overflow of hospital capacity. It is important to continue monitoring contacts as vaccines are rolled out.

## Data Availability

The datasets generated and/or analysed during the current study are not publicly available due to the dataset containing sensitive personal data. Aggregated data are available from the corresponding author on reasonable request.

## Acknowledgements

MCB acknowledges funding from the MRC Centre for Global Infectious Disease Analysis (reference MR/R015600/1), jointly funded by the UK Medical Research Council (MRC) and the UK Foreign, Commonwealth & Development Office (FCDO), under the MRC/FCDO Concordat agreement and is also part of the EDCTP2 programme supported by the European Union.

## Author’s contribution

MB, MD, and MCB designed the study. All authors (except AB) participated in the development and validation of the study questionnaires. MB and MD drafted the article and supervised the data collection and analysis. AG, GB, and LDR participated in data collection. AG, MM, GB, LDR, PLM, AB and ED participated in the analysis. All authors interpreted the results and critically revised the manuscript for scientific content. All authors approved the final version of the article.

## Funding

This study was funded by the Canadian Immunization Research Network, the Canadian Institutes of Health Research (foundation scheme grant FDN-143283), the Institut National de Santé Publique du Québec, and the Fonds de recherche du Québec – Santé research (scholars award to MB).

## Availability of data and materials

The datasets generated and/or analysed during the current study are not publicly available due to the dataset containing sensitive personal data. Aggregated data and mixing matrices data are available from the corresponding author on reasonable request.

## DECLARATIONS

### Ethics approval and consent to participate

All methods were carried out in accordance with relevant guidelines and regulations. The CONNECT study was approved by the Ethics Committee of the Centre de recherche du CHU de Québec-Université Laval (project 2016-2172). All participants provided informed consent during the recruitment phone call. Informed consent was taken from a parent and/or legal guardian for study participation in the case of minors.

### Consent for publication

Not applicable.

### Competing interests

The authors declare that they have no competing interests.

## Notes

### Competing Interest Statement

The authors have declared no competing interest.

### Author Declarations

The CONNECT study was approved by the Ethics Committee of the Centre de recherche du CHU de Quebec-Universite Laval (project 2016-2172).

### Summary of Updates

We have added a stringency index and the mixing matrices.

