## Supplementary material for "Time trends in social contacts before and during the COVID-19 pandemic: the CONNECT study"

#### Additional file 1

**Table S1. Overview of the different phases of CONNECT.**

| | **CONNECT1 Pre-COVID** | **CONNECT2 1st wave** | **CONNECT3,4,5 Summer 2020 and 2nd wave** |
|---|---|---|---|
| Data collection dates | Feb 1st 2018 – March 17th 2019 | April 21st 2020 – May 25th 2020 | July 3rd 2020 – February 26th 2021 |
| Funding | Canadian Immunization Network (CIRN) | Canadian Institute of Health Research (CIHR) Foundation Scheme Grant | Institut national de Santé publique du Québec (INSPQ) |
| Number of participants included in analysis | **Quebec: 1291** | **Quebec: 546** | **Quebec: 4970** |

**Table S2. Weighting procedure.**

New series of weights were calculated for each CONNECT phase (Pre-COVID, 1st wave, summer 2020 and 2nd wave) using multiple steps:

| Steps | Procedure |
|---|---|
| 1. Age and sex distribution | Weights W1 were calculated to replicate the sex-by-age distribution of the Quebec population, using the Canadian census data (8 age categories as presented in Table 1 by sex). |
| 2. Household composition | Weights W2 were calculated from W1 to replicate the Quebec household composition distribution in 4 categories, based on the age of youngest member (0-5, 6-14, 15-17, ≥ 18 years old, as presented in Table 1). We adjusted the weights to get close to the household composition without impacting the age distribution. |
| 3. Quebec regions | Weights W3 were calculated from W2 to replicate the distribution of Quebec regions (Greater Montreal*, Other regions). |
| 4. Final adjustment | Weights W4 were adjusted so that the sum of weights=n<br>We verified that W4-weighted distributions were generally representative of the Quebec population for key socio-demographic characteristics not considered in the weighting procedure (e.g. employment rate, ethnicity, mother tongue, country of origin) |
| 5. Days of the week | Weights W4 were multiplied by 5/7 or 2/7 for week or weekend days respectively, to obtain the daily number of social contacts on a weekly basis. |

* Greater Montreal: Regions of Montréal, Laval, Montérégie, Lanaudière, Laurentides.

#### Table S3. Time trends in the number of social contacts* by location of contacts.

##### A) Mean number of social contacts

| | Pre-COVID n=1291 | | Spring 2020 n=546 | | | Summer 2020 n=988 | | | September 2020 n=431 | | | Fall 2020 n=1052 | | | Holidays 2020-21 n=1569 | | | January 2021 n=501 | | | February 2021 n=429 | | |
|---|---|---|---|---|---|---|---|---|---|---|---|---|---|---|---|---|---|---|---|---|---|---|---|
| | **mean** | **95%CI** | **mean** | **95%CI** | **Diff† *p*** | **mean** | **95%CI** | **Diff† *p*** | **mean** | **95%CI** | **Diff† *p*** | **mean** | **95% CI** | **Diff† *p*** | **mean** | **95% CI** | **Diff† *p*** | **mean** | **95% CI** | **Diff† *p*** | **mean** | **95% CI** | **Diff† *p*** |
| Total | 7.8 | (7.2-8.5) | 3.1 | (2.6-3.5) | -4.8<br>*<.001* | 4.2 | (3.8-4.6) | 1.2<br>*<.001* | 5.0 | (4.3-5.8) | 0.8<br>*0.07* | 4.1 | (3.7-4.5) | -0.9<br>*0.04* | 2.9 | (2.7-3.1) | -1.2<br>*<.001* | 3.5 | (3.0-3.9) | 0.5<br>*0.02* | 4.0 | (3.3-4.6) | 0.5<br>*0.23* |
| Home (total) | 2.3 | (2.1-2.4) | 1.7 | (1.5-1.8) | -0.6<br>*<.001* | 2.3 | (2.1-2.4) | 0.6<br>*<.001* | 1.9 | (1.8-2.1) | -0.3<br>*0.004* | 1.7 | (1.7-1.8) | -0.2<br>*0.04* | 1.8 | (1.7-1.8) | 0.0<br>*0.75* | 1.7 | (1.5-1.8) | -0.1<br>*0.08* | 1.7 | (1.6-1.8) | 0.0<br>*0.58* |
| with household members | 1.2 | (1.1-1.3) | 1.3 | (1.2-1.5) | 0.1<br>*0.09* | 1.3 | (1.2-1.3) | -0.1<br>*0.30* | 1.3 | (1.2-1.4) | 0.1<br>*0.44* | 1.4 | (1.3-1.5) | 0.1<br>*0.46* | 1.4 | (1.3-1.4) | -0.0<br>*0.84* | 1.4 | (1.3-1.5) | 0.1<br>*0.24* | 1.4 | (1.2-1.5) | -0.1<br>*0.40* |
| with visitors | 1.1 | (1.0-1.2) | 0.3 | (0.2-0.4) | -0.8<br>*<.001* | 1.0 | (0.9-1.1) | 0.7<br>*<.001* | 0.6 | (0.5-0.7) | -0.4<br>*<.001* | 0.4 | (0.3-0.4) | -0.3<br>*<.001* | 0.4 | (0.4-0.4) | 0.0<br>*0.35* | 0.2 | (0.2-0.3) | -0.2<br>*<.001* | 0.3 | (0.3-0.4) | 0.1<br>*0.005* |
| Work£ | 2.7 | (2.1-3.2) | 1.2 | (0.8-1.6) | -1.5<br>*<.001* | 1.2 | (0.8-1.5) | -0.0<br>*0.99* | 1.5 | (0.9-2.2) | 0.4<br>*0.30* | 1.1 | (0.8-1.4) | -0.5<br>*0.18* | 0.8 | (0.6-1.1) | -0.3<br>*0.15* | 1.0 | (0.7-1.3) | 0.2<br>*0.43* | 0.9 | (0.5-1.4) | -0.1<br>*0.79* |
| School¶ | 1.6 | (1.2-2.0) | 0.0 | (0.0-0.0) | -1.6<br>*<.001* | 0.2 | (0.1-0.3) | 0.2<br>*<.001* | 0.9 | (0.6-1.3) | 0.7<br>*<.001* | 0.9 | (0.7-1.2) | -0.0<br>*0.94* | 0.0 | (0.0-0.1) | -0.9<br>*<.001* | 0.6 | (0.3-0.8) | 0.5<br>*<.001* | 1.0 | (0.6-1.5) | 0.5<br>*0.08* |
| Transport | 0.0 | (0.0-0.1) | 0.0 | (0.0-0.0) | -0.0<br>*<.001* | 0.0 | (0.0-0.0) | 0.0<br>*0.13* | 0.1 | (0.0-0.1) | 0.1<br>*0.03* | 0.0 | (0.0-0.0) | -0.0<br>*0.11* | 0.0 | (0.0-0.0) | -0.0<br>*0.003* | 0.0 | (0.0-0.0) | 0.0<br>*0.19* | 0.0 | (0.0-0.0) | -0.0<br>*0.62* |
| Leisure | 0.5 | (0.4-0.6) | 0.0 | (0.0-0.1) | -0.5<br>*<.001* | 0.2 | (0.2-0.3) | 0.2<br>*<.001* | 0.2 | (0.1-0.3) | -0.0<br>*0.28* | 0.1 | (0.0-0.1) | -0.1<br>*<.001* | 0.1 | (0.1-0.1) | 0.0<br>*0.03* | 0.1 | (0.1-0.1) | -0.0<br>*0.66* | 0.1 | (0.0-0.1) | -0.0<br>*0.76* |
| Other | 0.7 | (0.6-0.8) | 0.2 | (0.1-0.2) | -0.6<br>*<.001* | 0.3 | (0.3-0.4) | 0.2<br>*<.001* | 0.3 | (0.2-0.4) | -0.0<br>*0.35* | 0.2 | (0.2-0.3) | -0.0<br>*0.36* | 0.1 | (0.1-0.2) | -0.1<br>*<.001* | 0.1 | (0.1-0.2) | 0.0<br>*0.90* | 0.2 | (0.1-0.3) | 0.0<br>*0.30* |

**Pre-COVID:** February 1st 2018 to March 17th 2019**; Spring 2020**: April 21st to May 25th 2020; **Summer 2020**: July 3rd to August 31st 2020; **Fall 2020**: October 1st to December 16th 2020; **Holidays 2020-2021**: December 17th 2020 to January 8th 2021. **1st wave:** Spring 2020; **2nd wave**: September 2020 to February 2021.

*Mean number of social contacts weighted to represent daily contacts over one usual week, estimated at the population level.

† Diff: Absolute difference with the preceding period and p-value of this difference.

£: Work: Contacts at work are truncated to a maximum of 40 contacts per day.

¶ School: Contacts at school include contacts of workers in schools (truncated to a maximum of 40 per day).

#### B) Median number of social contacts

| | Pre-COVID n=1291 | | Spring 2020 n=546 | | Summer 2020 n=988 | | September 2020 n=431 | | Fall 2020 n=1052 | | Holidays 2020-21 n=1569 | | January 2021 n=501 | | February 2021 n=429 | |
|---|---|---|---|---|---|---|---|---|---|---|---|---|---|---|---|---|
| | **median** | **(Q1-Q3)** | **median** | **(Q1-Q3)** | **median** | **(Q1-Q3)** | **median** | **(Q1-Q3)** | **median** | **(Q1-Q3)** | **median** | **(Q1-Q3)** | **median** | **(Q1-Q3)** | **median** | **(Q1-Q3)** |
| Total | 4 | (2-9) | 2 | (1-3) | 3 | (1-5) | 2 | (1-5) | 2 | (1-4) | 2 | (1-3) | 2 | (1-4) | 2 | (1-4) |
| Home (total) | 2 | (1-3) | 1 | (0-3) | 2 | (1-3) | 1 | (1-3) | 1 | (1-3) | 1 | (1-3) | 1 | (1-3) | 1 | (1-3) |
| with household members | 1 | (0-2) | 1 | (0-2) | 1 | (0-2) | 1 | (0-2) | 1 | (0-2) | 1 | (0-2) | 1 | (0-2) | 1 | (0-2) |
| with visitors | 0 | (0-1) | 0 | (0-0) | 0 | (0-1) | 0 | (0-1) | 0 | (0-0) | 0 | (0-0) | 0 | (0-0) | 0 | (0-0) |
| Work[£] | 0 | (0-0) | 0 | (0-0) | 0 | (0-0) | 0 | (0-0) | 0 | (0-0) | 0 | (0-0) | 0 | (0-0) | 0 | (0-0) |
| School[¶] | 0 | (0-0) | 0 | (0-0) | 0 | (0-0) | 0 | (0-0) | 0 | (0-0) | 0 | (0-0) | 0 | (0-0) | 0 | (0-0) |
| Transport | 0 | (0-0) | 0 | (0-0) | 0 | (0-0) | 0 | (0-0) | 0 | (0-0) | 0 | (0-0) | 0 | (0-0) | 0 | (0-0) |
| Leisure | 0 | (0-0) | 0 | (0-0) | 0 | (0-0) | 0 | (0-0) | 0 | (0-0) | 0 | (0-0) | 0 | (0-0) | 0 | (0-0) |
| Other | 0 | (0-1) | 0 | (0-0) | 0 | (0-0) | 0 | (0-0) | 0 | (0-0) | 0 | (0-0) | 0 | (0-0) | 0 | (0-0) |

**Pre-COVID:** February 1st 2018 to March 17th 2019**; Spring 2020**: April 21st to May 25th 2020; **Summer 2020**: July 3rd to August 31st 2020; **Fall 2020**: October 1st to December 16th 2020; **Holidays 2020-2021**: December 17th 2020 to January 8th 2021. **1st wave:** Spring 2020; **2nd wave**: September 2020 to February 2021.

£: Work: Contacts at work are truncated to a maximum of 40 contacts per day.

¶ School: Contacts at school include contacts of workers in schools (truncated to a maximum of 40 per day).

**Table S4. Association between the mean number of social contacts and the stringency index.**

| | Stringency index[£] | | | | | | | | | | | | | |
|---|---|---|---|---|---|---|---|---|---|---|---|---|---|---|
| | **0 (Pre-COVID)** | | **40.0 - 49.9** | | | **50.0 - 59.9*** | | | **60.0 - 69.9*** | | | **≥ 70*** | | |
| | **mean** | **95%CI** | **mean** | **95%CI** | **Diff[†]**<br>***p*** | **mean** | **95%CI** | **Diff[†]**<br>***p*** | **mean** | **95%CI** | **Diff[†]**<br>***p*** | **mean** | **95% CI** | **Diff[†]**<br>***p*** |
| Total | 7.8 | (7.2-8.5) | 4.6 | (4.1-5.1) | -3.2<br>*<.0001* | 4.1 | (3.8-4.5) | -0.5<br>*0.09* | 3.5 | (3.2-3.8) | -0.6<br>*0.01* | 2.9 | (2.6-3.1) | -0.7<br>*0.0005* |
| Home (total) | 2.3 | (2.1-2.4) | 2.2 | (2.0-2.3) | -0.1<br>*0.23* | 1.9 | (1.8-2.0) | -0.3<br>*0.0002* | 1.7 | (1.6-1.7) | -0.2<br>*0.0008* | 1.8 | (1.7-1.8) | 0.1<br>*0.15* |
| with household members | 1.2 | (1.1-1.3) | 1.3 | (1.2-1.4) | 0.1<br>*0.13* | 1.3 | (1.3-1.4) | 0.0<br>*0.54* | 1.4 | (1.3-1.4) | 0.0<br>*0.57* | 1.4 | (1.3-1.5) | 0.0<br>*0.65* |
| with visitors | 1.1 | (1.0-1.2) | 0.9 | (0.8-0.9) | -0.2<br>*0.002* | 0.5 | (0.5-0.6) | -0.3<br>*<.0001* | 0.3 | (0.3-0.3) | -0.2<br>*<.0001* | 0.4 | (0.3-0.4) | 0.1<br>*0.03* |
| Work[£] | 2.7 | (2.1-3.2) | 1.3 | (0.9-1.7) | -1.3<br>*<.0001* | 1.1 | (0.9-1.4) | -0.2<br>*0.41* | 1.0 | (0.8-1.3) | -0.1<br>*0.54* | 0.9 | (0.7-1.1) | -0.2<br>*0.34* |
| School[¶] | 1.6 | (1.2-2.0) | 0.5 | (0.4-0.7) | -1.1<br>*<.0001* | 0.7 | (0.5-0.9) | 0.2<br>*0.14* | 0.5 | (0.4-0.7) | -0.2<br>*0.18* | 0.0 | (0.0-0.0) | -0.5<br>*<.0001* |
| Transport | 0.0 | (0.0-0.1) | 0.0 | (0.0-0.1) | 0.0<br>*0.57* | 0.0 | (0.0-0.0) | 0.0<br>*0.21* | 0.0 | (0.0-0.0) | 0.1<br>*0.02* | 0.0 | (0.0-0.0) | 0.0<br>*0.09* |
| Leisure | 0.5 | (0.4-0.6) | 0.2 | (0.2-0.3) | -0.3<br>*<.0001* | 0.1 | (0.1-0.1) | -0.1<br>*<.0001* | 0.1 | (0.1-0.1) | 0.0<br>*0.06* | 0.1 | (0.1-0.1) | 0.0<br>*0.99* |
| Other | 0.7 | (0.6-0.8) | 0.3 | (0.3-0.4) | -0.4<br>*<.0001* | 0.2 | (0.2-0.3) | -0.1<br>*0.04* | 0.2 | (0.2-0.2) | -0.1<br>*0.02* | 0.1 | (0.1-0.2) | -0.1<br>*0.006* |

[£] Stringency index: Higher values indicate stricter measures; the association between social contacts and the stringency index was examined irrespective of periods.

* For stringency index >50, the mean number of contacts for all locations was significantly lower than the pre-COVID period (stringency index = 0).

[†] Diff: Absolute difference with the preceding stringency index category and p-value of this difference.

[£:] Work: Contacts at work are truncated to a maximum of 40 contacts per day.

[¶] School: Contacts at school include contacts of workers in schools (truncated to a maximum of 40 per day).

**Table S5. Time trends in the total number of social contacts by key socio-demographic characteristics.**

| | Pre-COVID n=1291 | | Spring 2020 n=546 | | | Summer 2020 n=988 | | | September 2020 n=431 | | | Fall 2020 n=1052 | | | Holidays 2020-21 n=1569 | | | January 2021 n=501 | | | February 2021 n=429 | | |
|---|---|---|---|---|---|---|---|---|---|---|---|---|---|---|---|---|---|---|---|---|---|---|---|
| | **mean** | **95% CI** | **mean** | **95% CI** | **Diff† *p*** | **mean** | **95% CI** | **Diff† *p*** | **mean** | **95% CI** | **Diff† *p*** | **mean** | **95% CI** | **Diff† *p*** | **mean** | **95% CI** | **Diff† *p*** | **mean** | **95% CI** | **Diff† *p*** | **mean** | **95% CI** | **Diff† *p*** |
| Total | 7.8 | (7.2-8.5) | 3.1 | (2.6-3.5) | -4.8 | 4.2 | (3.8-4.6) | 1.2 | 5.0 | (4.3-5.8) | 0.8 | 4.1 | (3.7-4.5) | -0.9 | 2.9 | (2.7-3.1) | -1.2 | 3.5 | (3.0-3.9) | 0.5 | 4.0 | (3.3-4.6) | 0.5 |
| | | | | | *<.001* | | | *<.001* | | | *0.07* | | | *0.04* | | | *<.001* | | | *0.02* | | | *0.2* |
| Age | | | | | | | | | | | | | | | | | | | | | | | |
| 0-17 yrs old | 7.7 | (7.0-8.4) | 3.9 | (3.3-4.5) | -3.8 | 5.2 | (4.5-5.8) | 1.3 | 8.6 | (6.9-10.4) | 3.4 | 6.5 | (5.7-7.3) | -2.1 | 3.6 | (3.4-3.8) | -2.9 | 4.9 | (4.3-5.5) | 1.3 | 7.3 | (5.7-8.9) | 2.4 |
| | | | | | *<.001* | | | *0.005* | | | *<.001* | | | *0.03* | | | *<.001* | | | *<.001* | | | *0.007* |
| 18-65 yrs old | 9.0 | (8.0-10.0) | 3.3 | (2.6-3.9) | -5.7 | 4.4 | (3.8-4.9) | 1.1 | 4.8 | (3.8-5.8) | 0.5 | 4.1 | (3.6-4.5) | -0.8 | 3.1 | (2.7-3.4) | -1.0 | 3.6 | (3.0-4.2) | 0.5 | 3.3 | (2.5-4.2) | -0.3 |
| | | | | | *<.001* | | | *0.02* | | | *0.43* | | | *0.18* | | | *0.001* | | | *0.12* | | | *0.62* |
| ≥ 65 yrs old | 3.5 | (3.0-4.0) | 1.2 | (0.7-1.6) | -2.4 | 2.8 | (2.0-3.6) | 1.6 | 1.6 | (1.2-2.0) | -1.1 | 1.4 | (1.1-1.6) | -0.3 | 1.2 | (1.0-1.5) | -0.1 | 1.4 | (0.8-1.9) | 0.1 | 1.6 | (1.2-2.0) | 0.3 |
| | | | | | *<.001* | | | *<.001* | | | *0.02* | | | *0.29* | | | *0.48* | | | *0.67* | | | *0.44* |
| *p-value*¥ | | *<0.001* | | *<0.001* | | | *<0.001* | | | *<0.001* | | | *<0.001* | | | *<0.001* | | | *<0.001* | | | *<0.001* | |
| Sex | | | | | | | | | | | | | | | | | | | | | | | |
| Male | 7.4 | (6.4-8.3) | 2.9 | (2.5-3.3) | -4.4 | 3.9 | (3.3-4.4) | 1.0 | 4.5 | (3.8-5.2) | 0.6 | 3.9 | (3.4-4.4) | -0.6 | 3.2 | (2.9-3.5) | -0.7 | 3.6 | (3.1-4.1) | 0.4 | 3.7 | (3.0-4.4) | 0.1 |
| | | | | | *<.001* | | | *0.005* | | | *0.18* | | | *0.18* | | | *0.02* | | | *0.12* | | | *0.74* |
| Female | 8.1 | (7.3-8.8) | 3.5 | (2.9-4.0) | -4.6 | 4.8 | (4.2-5.3) | 1.3 | 6.2 | (4.5-7.8) | 1.4 | 4.1 | (3.7-4.5) | -2.1 | 3.0 | (2.7-3.2) | -1.1 | 3.6 | (3.0-4.2) | 0.6 | 3.6 | (2.9-4.3) | 0.0 |
| | | | | | *<.001* | | | *0.001* | | | *0.12* | | | *0.02* | | | *<.001* | | | *0.06* | | | *0.92* |
| *p-value*¥ | | *0.26* | | *0.12* | | | *0.02* | | | *0.06* | | | *0.53* | | | *0.21* | | | *0.99* | | | *0.85* | |
| Region | | | | | | | | | | | | | | | | | | | | | | | |
| Urban | 7.5 | (6.9-8.1) | 3.2 | (2.8-3.6) | -4.4 | 4.4 | (3.9-4.8) | 1.2 | 5.1 | (4.3-5.8) | 0.7 | 4.0 | (3.6-4.4) | -1.1 | 3.1 | (2.8-3.3) | -0.9 | 3.5 | (3.1-3.8) | 0.4 | 3.7 | (3.1-4.2) | 0.2 |
| | | | | | *<.001* | | | *<.001* | | | *0.12* | | | *0.01* | | | *<.001* | | | *0.08* | | | *0.55* |
| Rural | 8.8 | (6.9-10.8) | 3.5 | (2.6-4.4) | -5.4 | 4.9 | (3.7-6.1) | 1.4 | 3.8 | (2.3-5.3) | -1.1 | 4.2 | (3.3-5.0) | 0.4 | 3.2 | (2.9-3.5) | -1.0 | 4.0 | (3.1-5.0) | 0.8 | 3.9 | (2.6-5.3) | -0.1 |
| | | | | | *<.001* | | | *0.06* | | | *0.26* | | | *0.67* | | | *0.03* | | | *0.10* | | | *0.92* |
| *p-value*¥ | | *0.19* | | *0.51* | | | *0.42* | | | *0.14* | | | *0.74* | | | *0.60* | | | *0.28* | | | *0.70* | |
| Region | | | | | | | | | | | | | | | | | | | | | | | |
| Greater Mtl€ | 8.0 | (7.0-8.9) | 2.9 | (2.4-3.5) | -5.0 | 3.9 | (3.5-4.3) | 0.9 | 4.8 | (3.9-5.6) | 0.9 | 3.9 | (3.4-4.3) | -0.9 | 2.9 | (2.5-3.3) | -0.9 | 3.4 | (2.7-4.0) | 0.5 | 3.8 | (2.9-4.7) | 0.4 |
| | | | | | *<.001* | | | *0.006* | | | *0.06* | | | *0.07* | | | *0.001* | | | *0.21* | | | *0.49* |
| Other regions | 7.1 | (6.3-8.0) | 3.2 | (2.3-4.1) | -4.0 | 4.5 | (3.6-5.4) | 1.4 | 5.0 | (3.5-6.4) | 0.4 | 4.4 | (3.6-5.2) | -0.5 | 2.7 | (2.5-3.0) | -1.7 | 3.4 | (2.8-3.9) | 0.6 | 3.8 | (2.9-4.6) | 0.4 |
| | | | | | *<.001* | | | *0.03* | | | *0.63* | | | *0.52* | | | *<.001* | | | *0.05* | | | *0.43* |
| *p-value*¥ | | *0.19* | | *0.67* | | | *0.17* | | | *0.85* | | | *0.26* | | | *0.46* | | | *0.96* | | | *0.99* | |
| Material deprivation | | | | | | | | | | | | | | | | | | | | | | | |
| Q1-Most privileged | 7.3 | (6.3-8.2) | 2.8 | (2.3-3.2) | -4.5 | 4.9 | (4.1-5.6) | 2.1 | 5.2 | (3.9-6.6) | 0.4 | 3.6 | (3.1-4.1) | -1.7 | 3.0 | (2.7-3.3) | -0.6 | 3.2 | (2.4-3.9) | 0.2 | 3.2 | (2.7-3.8) | 0.0 |
| | | | | | *<.001* | | | *<.001* | | | *0.62* | | | *0.02* | | | *0.02* | | | *0.60* | | | *0.92* |
| Q2 | 7.2 | (6.0-8.4) | 2.9 | (2.0-3.8) | -4.3 | 4.1 | (3.4-4.7) | 1.2 | 3.9 | (3.1-4.8) | -0.1 | 4.0 | (3.3-4.6) | 0.0 | 3.5 | (2.8-4.2) | -0.5 | 3.6 | (2.8-4.3) | 0.1 | 3.8 | (2.9-4.6) | 0.2 |
| | | | | | *<.001* | | | *0.04* | | | *0.85* | | | *0.99* | | | *0.36* | | | *0.90* | | | *0.69* |
| Q3 | 8.1 | (6.6-9.7) | 3.4 | (2.5-4.2) | -4.8 | 4.1 | (3.3-5.0) | 0.8 | 6.6 | (4.1-9.1) | 2.5 | 4.4 | (3.5-5.3) | -2.2 | 3.2 | (2.8-3.7) | -1.2 | 3.3 | (2.8-3.9) | 0.1 | 3.6 | (2.4-4.8) | 0.3 |
| | | | | | *<.001* | | | *0.20* | | | *0.07* | | | *0.10* | | | *0.03* | | | *0.79* | | | *0.68* |
| Q4 | 7.7 | (6.4-9.0) | 3.9 | (2.8-5.0) | -3.8 | 4.1 | (3.2-4.9) | 0.2 | 5.0 | (3.7-6.4) | 1.0 | 4.3 | (3.5-5.2) | -0.7 | 2.9 | (2.6-3.1) | -1.5 | 4.2 | (3.3-5.1) | 1.4 | 4.1 | (2.8-5.5) | -0.1 |
| | | | | | *<.001* | | | *0.79* | | | *0.22* | | | *0.37* | | | *<.001* | | | *0.003* | | | *0.93* |
| Q5-Most deprived | 8.5 | (6.5-10.4) | 3.3 | (2.4-4.1) | -5.2 | 4.5 | (3.3-5.6) | 1.2 | 3.3 | (1.7-5.0) | -1.1 | 4.1 | (2.9-5.2) | 0.8 | 3.0 | (2.6-3.4) | -1.1 | 3.4 | (2.3-4.4) | 0.4 | 4.1 | (1.9-6.4) | 0.7 |
| | | | | | *<.001* | | | *0.11* | | | *0.27* | | | *0.45* | | | *0.08* | | | *0.51* | | | *0.56* |
| *p-value*¥ | | *0.72* | | *0.26* | | | *0.59* | | | *0.09* | | | *0.40* | | | *0.41* | | | *0.44* | | | *0.64* | |

| | Pre-COVID n=1291 | | Spring 2020 n=546 | | | Summer 2020 n=988 | | | September 2020 n=431 | | | Fall 2020 n=1052 | | | Holidays 2020-21 n=1569 | | | January 2021 n=501 | | | February 2021 n=429 | | |
|---|---|---|---|---|---|---|---|---|---|---|---|---|---|---|---|---|---|---|---|---|---|---|---|
| | **mean** | **95% CI** | **mean** | **95% CI** | **Diff[†]**<br>***p*** | **mean** | **95% CI** | **Diff[†]**<br>***p*** | **mean** | **95% CI** | **Diff[†]**<br>***p*** | **mean** | **95% CI** | **Diff[†]**<br>***p*** | **mean** | **95% CI** | **Diff[†]**<br>***p*** | **mean** | **95% CI** | **Diff[†]**<br>***p*** | **mean** | **95% CI** | **Diff[†]**<br>***p*** |
| Social deprivation | | | | | | | | | | | | | | | | | | | | | | | |
| Q1-Most privileged | 7.7 | (6.1-9.3) | 3.7 | (2.5-4.8) | -4.1<br>*<.001* | 4.7 | (3.7-5.8) | 1.1<br>*0.18* | 6.0 | (3.8-8.3) | 1.3<br>*0.31* | 3.8 | (3.2-4.4) | -2.3<br>*0.06* | 3.3 | (2.8-3.9) | -0.4<br>*0.28* | 4.2 | (3.1-5.4) | 0.9<br>*0.17* | 4.4 | (2.9-5.9) | 0.1<br>*0.90* |
| Q2 | 8.4 | (6.8-9.9) | 3.3 | (2.3-4.3) | -5.1<br>*<.001* | 4.3 | (3.6-5.0) | 1.0<br>*0.11* | 4.0 | (3.1-4.9) | -0.2<br>*0.67* | 4.1 | (3.4-4.7) | 0.1<br>*0.91* | 3.2 | (2.8-3.7) | -0.8<br>*0.04* | 3.9 | (2.9-5.0) | 0.7<br>*0.22* | 3.2 | (2.5-3.9) | -0.8<br>*0.23* |
| Q3 | 7.4 | (6.2-8.6) | 3.0 | (2.3-3.8) | -4.4<br>*<.001* | 4.1 | (3.5-4.8) | 1.1<br>*0.03* | 5.2 | (3.7-6.7) | 1.1<br>*0.19* | 4.4 | (3.5-5.3) | -0.8<br>*0.34* | 3.4 | (2.8-3.9) | -1.0<br>*0.06* | 3.6 | (2.9-4.3) | 0.2<br>*0.60* | 4.1 | (2.6-5.6) | 0.5<br>*0.58* |
| Q4 | 6.9 | (5.7-8.1) | 3.4 | (2.4-4.3) | -3.5<br>*<.001* | 4.2 | (3.7-4.7) | 0.9<br>*0.11* | 5.5 | (3.8-7.2) | 1.3<br>*0.17* | 4.4 | (3.5-5.3) | -1.1<br>*0.28* | 3.0 | (2.6-3.4) | -1.4<br>*0.004* | 3.1 | (2.7-3.6) | 0.1<br>*0.62* | 3.6 | (2.9-4.4) | 0.5<br>*0.26* |
| Q5-Most deprived | 8.1 | (6.8-9.4) | 2.9 | (2.3-3.5) | -5.2<br>*<.001* | 4.6 | (3.4-5.9) | 1.7<br>*0.02* | 4.1 | (3.0-5.3) | -0.5<br>*0.55* | 3.6 | (2.9-4.2) | -0.6<br>*0.38* | 2.7 | (2.4-3.1) | -0.8<br>*0.02* | 3.2 | (2.5-3.9) | 0.5<br>*0.20* | 3.5 | (2.4-4.6) | 0.3<br>*0.63* |
| *p-value[¥]* | | *0.54* | | *0.80* | | | *0.85* | | | *0.25* | | | *0.42* | | | *0.16* | | | *0.29* | | | *0.58* | |
| Level of education* | | | | | | | | | | | | | | | | | | | | | | | |
| No diploma | 6.0 | (3.2-8.8) | 4.1 | (0.7-7.5) | -1.9<br>*0.41* | 7.2 | (0.0-15.4) | 3.1<br>*0.49* | 3.3 | (0.0-7.1) | -3.9<br>*0.34* | 6.8 | (1.6-12.0) | 3.5<br>*0.28* | 3.4 | (0.9-6.0) | -3.4<br>*0.26* | 2.2 | (1.7-2.8) | -1.2<br>*0.37* | 1.5 | (0.9-2.2) | -0.7<br>*0.11* |
| High school | 8.8 | (5.6-12.0) | 5.5 | (2.8-8.3) | -3.2<br>*0.13* | 3.6 | (2.1-5.1) | -2.0<br>*0.22* | 5.9 | (1.8-10.1) | 2.4<br>*0.28* | 4.2 | (2.4-5.9) | -1.8<br>*0.44* | 2.5 | (1.8-3.2) | -1.6<br>*0.09* | 4.2 | (2.2-6.2) | 1.7<br>*0.12* | 4.5 | (1.1-8.0) | 0.3<br>*0.87* |
| College, cegep,non-university | 8.4 | (6.7-10.0) | 3.3 | (2.1-4.4) | -5.1<br>*<.001* | 4.2 | (3.0-5.3) | 0.9<br>*0.27* | 5.7 | (3.0-8.4) | 1.6<br>*0.29* | 4.3 | (3.3-5.4) | -1.4<br>*0.33* | 3.2 | (2.6-3.8) | -1.1<br>*0.07* | 4.4 | (3.0-5.8) | 1.2<br>*0.11* | 3.0 | (1.9-4.2) | -1.4<br>*0.14* |
| University | 10.7 | (9.0-12.5) | 2.5 | (1.7-3.3) | -8.2<br>*<.001* | 4.0 | (3.4-4.6) | 1.5<br>*0.003* | 4.4 | (2.9-5.8) | 0.4<br>*0.62* | 4.0 | (3.3-4.8) | -0.4<br>*0.66* | 3.1 | (2.5-3.7) | -0.9<br>*0.05* | 3.5 | (2.5-4.5) | 0.4<br>*0.47* | 2.8 | (2.0-3.7) | -0.7<br>*0.31* |
| *p-value[¥]* | | *0.03* | | *0.15* | | | *0.80* | | | *0.66* | | | *0.76* | | | *0.52* | | | *0.006* | | | *0.02* | |
| Household size | | | | | | | | | | | | | | | | | | | | | | | |
| 1-2 | 6.7 | (6.0-7.4) | 2.6 | (2.1-3.1) | -4.1<br>*<.001* | 3.8 | (3.2-4.3) | 1.2<br>*<.001* | 3.7 | (3.0-4.5) | -0.0<br>*0.95* | 3.2 | (2.8-3.7) | -0.5<br>*0.22* | 2.4 | (2.1-2.7) | -0.8<br>*<.001* | 2.9 | (2.5-3.4) | 0.5<br>*0.03* | 2.4 | (2.0-2.8) | -0.6<br>*0.06* |
| 3-4 | 9.1 | (7.7-10.5) | 3.7 | (2.9-4.5) | -5.4<br>*<.001* | 4.7 | (4.1-5.3) | 1.0<br>*0.05* | 6.1 | (4.6-7.6) | 1.4<br>*0.09* | 4.9 | (4.2-5.6) | -1.2<br>*0.16* | 3.6 | (3.0-4.1) | -1.3<br>*0.001* | 4.2 | (3.4-5.0) | 0.6<br>*0.14* | 6.0 | (4.5-7.6) | 1.8<br>*0.04* |
| ≥ 5 | 9.4 | (6.8-11.9) | 4.4 | (3.4-5.5) | -4.9<br>*<.001* | 7.8 | (5.9-9.7) | 3.4<br>*0.002* | 9.2 | (6.7-11.7) | 1.4<br>*0.38* | 6.7 | (5.5-7.9) | -2.5<br>*0.08* | 5.3 | (4.3-6.3) | -1.4<br>*0.07* | 4.9 | (4.1-5.7) | -0.4<br>*0.54* | 7.8 | (4.6-11.0) | 2.9<br>*0.08* |
| *p-value[¥]* | | *0.004* | | *0.003* | | | *<.001* | | | *<.001* | | | *<.001* | | | *<.001* | | | *<.001* | | | *<.001* | |
| Household composition | | | | | | | | | | | | | | | | | | | | | | | |
| With 0-17-yr-olds | 9.6 | (8.0-11.1) | 4.1 | (3.3-4.9) | -5.5<br>*<.001* | 5.5 | (4.7-6.3) | 1.4<br>*0.005* | 7.4 | (5.9-8.8) | 1.9<br>*0.02* | 6.1 | (5.3-6.9) | -1.3<br>*0.11* | 3.9 | (3.3-4.5) | -2.1<br>*<.001* | 4.3 | (3.7-4.9) | 0.4<br>*0.25* | 7.2 | (5.6-8.7) | 2.9<br>*<.001* |
| Without 0-17-yr-olds | 6.8 | (6.1-7.4) | 2.6 | (2.1-3.1) | -4.2<br>*<.001* | 3.8 | (3.2-4.3) | 1.2<br>*<.001* | 3.7 | (2.9-4.4) | -0.1<br>*0.84* | 3.1 | (2.7-3.5) | -0.6<br>*0.12* | 2.5 | (2.1-2.8) | -0.6<br>*0.002* | 3.1 | (2.5-3.6) | 0.6<br>*0.02* | 2.3 | (1.9-2.8) | -0.7<br>*0.02* |
| *p-value[¥]* | | *0.002* | | *0.004* | | | *0.001* | | | *<.001* | | | *<.001* | | | *<.001* | | | *0.004* | | | *<.001* | |
| Race | | | | | | | | | | | | | | | | | | | | | | | |
| Caucasian | 7.8 | (7.2-8.5) | 3.3 | (2.9-3.7) | -4.5<br>*<.001* | 4.6 | (4.1-5.0) | 1.3<br>*<.001* | 4.9 | (4.2-5.7) | 0.4<br>*0.39* | 4.1 | (3.7-4.4) | -0.9<br>*0.04* | 3.1 | (2.9-3.4) | -0.9<br>*<.001* | 3.7 | (3.3-4.1) | 0.5<br>*0.02* | 3.7 | (3.2-4.2) | 0.0<br>*0.91* |
| Other | 6.9 | (4.9-8.8) | 2.8 | (1.6-4.1) | -4.1<br>*<.001* | 3.1 | (2.2-4.0) | 0.3<br>*0.74* | 4.6 | (2.7-6.6) | 1.5<br>*0.16* | 3.6 | (2.6-4.6) | -1.0<br>*0.36* | 2.9 | (2.0-3.8) | -0.7<br>*0.34* | 2.7 | (1.9-3.5) | -0.2<br>*0.72* | 3.9 | (0.7-7.2) | 1.2<br>*0.47* |
| *p-value[¥]* | | *0.37* | | *0.50* | | | *0.005* | | | *0.77* | | | *0.38* | | | *0.64* | | | *0.04* | | | *0.89* | |

| | Pre-COVID n=1291 | | Spring 2020 n=546 | | | Summer 2020 n=988 | | | September 2020 n=431 | | | Fall 2020 n=1052 | | | Holidays 2020-21 n=1569 | | | January 2021 n=501 | | | February 2021 n=429 | | |
|---|---|---|---|---|---|---|---|---|---|---|---|---|---|---|---|---|---|---|---|---|---|---|---|
| | **mean** | **95% CI** | **mean** | **95% CI** | **Diff[†] *p*** | **mean** | **95% CI** | **Diff[†] *p*** | **mean** | **95% CI** | **Diff[†] *p*** | **mean** | **95% CI** | **Diff[†] *p*** | **mean** | **95% CI** | **Diff[†] *p*** | **mean** | **95% CI** | **Diff[†] *p*** | **mean** | **95% CI** | **Diff[†] *p*** |
| Country of origin | | | | | | | | | | | | | | | | | | | | | | | |
| Canada | 7.7 | (7.1-8.4) | 3.2 | (2.9-3.6) | -4.5 | 4.4 | (4.0-4.8) | 1.2 | 4.9 | (4.2-5.7) | 0.5 | 4.1 | (3.7-4.4) | -0.9 | 3.1 | (2.9-3.3) | -1.0 | 3.7 | (3.3-4.1) | 0.6 | 3.6 | (3.1-4.1) | -0.0 |
| | | | | | *<.001* | | | *<.001* | | | *0.19* | | | *0.04* | | | *<.001* | | | *0.006* | | | *0.95* |
| Other | 7.5 | (5.5-9.4) | 2.6 | (1.5-3.7) | -4.8 | 4.7 | (2.3-7.1) | 2.1 | 4.3 | (2.6-6.1) | -0.4 | 3.6 | (2.9-4.4) | -0.7 | 3.5 | (2.3-4.7) | -0.1 | 2.9 | (2.3-3.6) | -0.6 | 3.7 | (1.5-6.0) | 0.8 |
| | | | | | *<.001* | | | *0.12* | | | *0.81* | | | *0.48* | | | *0.86* | | | *0.39* | | | *0.48* |
| *p-value*[¥] | | *0.79* | | *0.28* | | | *0.81* | | | *0.53* | | | *0.28* | | | *0.46* | | | *0.06* | | | *0.92* | |
| Mother tongue | | | | | | | | | | | | | | | | | | | | | | | |
| French | 7.9 | (7.2-8.6) | 3.3 | (2.9-3.7) | -4.6 | 4.5 | (4.0-5.0) | 1.2 | 4.9 | (4.1-5.6) | 0.4 | 4.1 | (3.7-4.4) | -0.8 | 3.1 | (2.9-3.3) | -1.0 | 3.7 | (3.3-4.1) | 0.6 | 3.9 | (3.3-4.5) | 0.2 |
| | | | | | *<.001* | | | *<.001* | | | *0.42* | | | *0.07* | | | *<.001* | | | *0.009* | | | *0.59* |
| English | 7.4 | (5.7-9.0) | 2.2 | (1.5-2.9) | -5.2 | 4.9 | (3.7-6.0) | 2.7 | 4.8 | (3.1-6.5) | -0.1 | 4.0 | (3.0-5.1) | -0.7 | 2.6 | (2.1-3.2) | -1.4 | 3.3 | (2.3-4.4) | 0.7 | 2.4 | (1.4-3.4) | -0.9 |
| | | | | | *<.001* | | | *<.001* | | | *0.90* | | | *0.47* | | | *0.02* | | | *0.25* | | | *0.19* |
| Other | 5.2 | (3.3-7.1) | 2.4 | (1.4-3.5) | -2.8 | 2.5 | (2.2-2.8) | 0.1 | 5.2 | (2.5-7.8) | 2.6 | 3.6 | (2.7-4.4) | -1.6 | 3.8 | (2.0-5.6) | 0.2 | 2.6 | (2.1-3.2) | -1.1 | 2.7 | (1.3-4.1) | 0.1 |
| | | | | | *0.01* | | | *0.88* | | | *0.05* | | | *0.26* | | | *0.83* | | | *0.23* | | | *0.91* |
| *p-value*[¥] | | *0.03* | | *0.02* | | | *<.001* | | | *0.97* | | | *0.54* | | | *0.25* | | | *0.006* | | | *0.03* | |

**Pre-COVID:** February 1st 2018 to March 17th 2019**; Spring 2020**: April 21st to May 25th 2020; **Summer 2020**: July 3rd to August 31st 2020; **Fall 2020**: October 1st to December 16th 2020; **Holidays 2020-2021**: December 17th 2020 to January 8th 2021. **1st wave:** Spring 2020; **2nd wave**: September 2020 to February 2021.

Comparisons are adjusted for age, except for the total estimate, estimates stratified by age, and estimates stratified by Greater Montréal and Other regions (Greater Montréal and Other regions are weighted to represent their respective age distribution).

* Among participants aged 25-64 years.

[†] Diff: Absolute difference with the preceding period and p-value of this difference

[¥] p-value of difference between the categories of a given characteristic for each time point.

[€] Greater Mtl: Regions of Montréal, Laval, Montérégie, Lanaudière, Laurentides.

**Table S6. Time trends in the number of social contacts* at school/daycare among children-students according to school level.**

| | Pre-COVID School season n=427 | | Spring 2020 n=167 | | | Summer 2020 n=250 | | | September 2020 n=119 | | | Fall 2020 n=293 | | | Holidays 2020-21 n=442 | | | January 2021 n=145 | | | February 2021 n=112 | | |
|---|---|---|---|---|---|---|---|---|---|---|---|---|---|---|---|---|---|---|---|---|---|---|---|
| | **mean** | **95% CI** | **mean** | **95% CI** | **Diff† p** | **mean** | **95% CI** | **Diff† p** | **mean** | **95% CI** | **Diff† p** | **mean** | **95% CI** | **Diff† p** | **mean** | **95% CI** | **Diff† p** | **mean** | **95% CI** | **Diff† p** | **mean** | **95% CI** | **Diff† p** |
| All students | 3.1 | (2.5-3.6) | 0.0 | (0.0-0.0) | -3.0<br>*<.001* | 0.6 | (0.3-1.0) | 0.6<br>*<.001* | 2.9 | (1.9-4.0) | 2.3<br>*<.001* | 2.0 | (1.5-2.5) | -0.9<br>*0.14* | 0.1 | (0.0-0.2) | -1.9<br>*<.001* | 1.1 | (0.6-1.5) | 1.0<br>*<.001* | 3.0 | (1.8-4.2) | 1.9<br>*0.004* |
| Daycare | 3.6 | (3.1-4.2) | 0.0 | (0.0-0.1) | -3.6<br>*<.001* | 1.9 | (0.7-3.1) | 1.9<br>*0.002* | 2.8 | (0.7-4.8) | 0.8<br>*0.49* | 4.2 | (2.9-5.5) | 1.4<br>*0.24* | 0.3 | (0.1-0.6) | -3.9<br>*<.001* | 3.3 | (1.8-4.8) | 3.0<br>*<.001* | 2.4 | (0.9-4.0) | -0.8<br>*0.46* |
| Elementary school | 3.8 | (2.9-4.7) | 0.0 | NA | -3.8<br>*NA* | 0.2 | (0.0-0.5) | 0.2<br>*NA* | 6.3 | (2.5-10.1) | 6.0<br>*0.002* | 5.2 | (3.2-7.2) | -1.1<br>*0.63* | 0.0 | (0.0-0.0) | -5.2<br>*<.001* | 1.5 | (0.4-2.7) | 1.5<br>*0.008* | 5.9 | (2.7-9.1) | 4.3<br>*0.01* |
| High school | 4.2 | (2.8-5.7) | 0.0 | NA | -4.2<br>*NA* | 0.0 | (0.0-0.1) | 0.0<br>*NA* | 4.2 | (2.3-6.0) | 4.1<br>*<.001* | 1.8 | (1.2-2.4) | -2.4<br>*0.02* | 0.0 | (0.0-0.0) | -1.8<br>*<.001* | 0.5 | (0.1-0.8) | 0.5<br>*0.007* | 3.2 | (1.5-5.0) | 2.7<br>*0.003* |
| Post-secondary | 1.2 | (0.6-1.7) | 0.0 | NA | -1.2<br>*NA* | 0.4 | (0.0-1.1) | 0.4<br>*NA* | 0.1 | (0.0-0.2) | -0.3<br>*0.30* | 0.1 | (0.0-0.2) | 0.0<br>*0.80* | 0.1 | (0.0-0.2) | -0.0<br>*0.94* | 0.0 | (0.0-0.1) | -0.0<br>*0.67* | 0.1 | (0.0-0.2) | 0.0<br>*0.64* |

**Pre-COVID school season:** February 1st 2018 to March 17th 2019, excluding July and August 2018**; Spring 2020**: April 21st to May 25th 2020; **Summer 2020**: July 3rd to August 31st 2020; **Fall 2020**: October 1st to December 16th 2020; **Holidays 2020-2021**: December 17th 2020 to January 8th 2021. **1st wave:** Spring 2020; **2nd wave**: September 2020 to February 2021.

NA: Not available because there was 0 contact during Spring 2020 (all schools closed).

***** Mean number of social contacts at school weighted to represent the mean daily contact over one usual week, estimated among students only.

† Diff: Absolute difference with the preceding period and p-value of this difference.

The mean numbers of contacts in this table are not exactly the same as those for school in Figure 2A because 1) the pre-COVID estimates are restricted to the school season, 2) the estimates include all students, irrespective of their age, 3) the mean is estimated among students only whereas in Figure 2A the mean is estimated among all 0-17-year-olds, some of whom do not attend school.

**Table S7. Time trends in social contacts at work* among workers and according to the type of employment (2016 National occupation classification).**

| | Pre-COVID n=355 | | Spring 2020 n=272 | | | Summer 2020 n=515 | | | September 2020 n=231 | | | Fall 2020 n=531 | | | Holidays 2020-21 n=778 | | | January 2021 n=251 | | | February 2021 n=214 | | |
|---|---|---|---|---|---|---|---|---|---|---|---|---|---|---|---|---|---|---|---|---|---|---|---|
| | **mean** | **95% CI** | **mean** | **95% CI** | **Diff† *p*** | **mean** | **95% CI** | **Diff† *p*** | **mean** | **95% CI** | **Diff † *p*** | **mean** | **95% CI** | **Diff† *p*** | **mea n** | **95% CI** | **Diff† *p*** | **mean** | **95% CI** | **Diff† *p*** | **mean** | **95% CI** | **Diff† *p*** |
| All workers | 7.3 | (6.1-8.6) | 2.6 | (1.7-3.4) | -4.8<br>*<.001* | 2.5 | (1.7-3.2) | -0.1<br>*0.85* | 3.0 | (1.8-4.1) | 0.5<br>*0.46* | 2.4 | (1.9-3.0) | -0.5<br>*0.42* | 1.8 | (1.3-2.3) | -0.6<br>*0.09* | 2.1 | (1.5-2.8) | 0.3<br>*0.43* | 1.9 | (1.0-2.9) | -0.2<br>*0.74* |
| 0. Management | 10.2 | (5.7-14.8) | 3.2 | (0.0-6.4) | -7.0<br>*0.01* | 4.7 | (1.1-8.3) | 1.5<br>*0.54* | 1.1 | (0.0-2.7) | -3.6<br>*0.07* | 2.0 | (0.7-3.2) | 0.8<br>*0.41* | 1.8 | (0.3-3.2) | -0.2<br>*0.83* | 2.3 | (0.3-4.2) | 0.5<br>*0.68* | 3.7 | (0.2-7.2) | 1.4<br>*0.49* |
| 1. Business, finance & administration | 6.5 | (4.3-8.7) | 0.5 | (0.1-0.9) | -6.0<br>*<.001* | 0.7 | (0.2-1.1) | 0.2<br>*0.46* | 2.9 | (0.6-5.2) | 2.2<br>*0.07* | 0.6 | (0.3-0.9) | -2.2<br>*0.06* | 0.4 | (0.1-0.6) | -0.2<br>*0.26* | 0.6 | (0.0-1.4) | 0.3<br>*0.50* | 1.2 | (0.0-2.8) | 0.5<br>*0.57* |
| 2. Natural & applied sciences | 3.4 | (1.9-5.0) | 0.2 | (0.0-0.4) | -3.3<br>*<.001* | 0.4 | (0.1-0.7) | 0.3<br>*0.15* | 0.5 | (0.0-1.0) | 0.1<br>*0.85* | 0.7 | (0.2-1.2) | 0.2<br>*0.57* | 0.4 | (0.1-0.8) | -0.3<br>*0.44* | 1.1 | (0.0-2.6) | 0.6<br>*0.43* | 0.4 | (0.1-0.8) | -0.6<br>*0.43* |
| 3. Health | 10.1 | (6.8-13.5) | 6.7 | (2.6-10.8) | -3.4<br>*0.20* | 4.2 | (2.2-6.2) | -2.5<br>*0.27* | 5.0 | (0.0-11.2) | 0.8<br>*0.81* | 7.9 | (4.8-10.9) | 2.9<br>*0.42* | 7.3 | (4.4-10.2) | -0.5<br>*0.80* | 7.3 | (3.6-11.1) | 0.0<br>*0.99* | 2.9 | (0.0-6.1) | -4.5<br>*0.07* |
| 4. Education, law &social, community & gov.services | 4.0 | (1.9-6.0) | 1.6 | (0.0-4.1) | -2.4<br>*0.15* | 0.5 | (0.2-0.9) | -1.0<br>*0.43* | 1.2 | (0.0-3.2) | 0.7<br>*0.50* | 1.0 | (0.4-1.6) | -0.2<br>*0.85* | 0.9 | (0.0-1.8) | -0.2<br>*0.79* | 0.1 | (0.0-0.3) | -0.7<br>*0.15* | 0.7 | (0.0-1.5) | 0.6<br>*0.13* |
| 5. Art, culture, recreation & sport | 0.7 | (0.1-1.3) | 0.1 | (0.0-0.3) | -0.6<br>*0.09* | 2.2 | (0.2-4.2) | 2.1<br>*0.05* | 1.8 | (0.0-4.6) | -0.3<br>*0.84* | 0.4 | (0.0-0.9) | -1.4<br>*0.32* | 0.4 | (0.0-1.0) | -0.1<br>*0.90* | NA | | | NA | | |
| 6. Sales & services | 10.8 | (7.9-13.7) | 4.7 | (1.7-7.7) | -6.1<br>*0.005* | 7.4 | (3.9-11.0) | 2.7<br>*0.25* | 8.4 | (3.5-13.4) | 1.0<br>*0.75* | 4.9 | (2.5-7.3) | -3.5<br>*0.21* | 3.9 | (2.0-5.8) | -1.1<br>*0.49* | 3.2 | (1.2-5.2) | -0.7<br>*0.61* | 5.6 | (1.3-9.8) | 2.4<br>*0.32* |
| 7. Trades, transport & equipment operators | 3.4 | (1.4-5.3) | 3.3 | (0.5-6.0) | -0.1<br>*0.95* | 1.6 | (0.5-2.7) | -1.7<br>*0.26* | 1.5 | (0.1-2.9) | -0.1<br>*0.93* | 3.1 | (0.9-5.4) | 1.6<br>*0.22* | 0.9 | (0.3-1.5) | -2.2<br>*0.06* | 1.0 | (0.1-1.9) | 0.1<br>*0.85* | 1.1 | (0.0-2.6) | 0.1<br>*0.92* |
| 8. Natural resources, agriculture & related production | 6.0 | (0.0-16.5) | 4.3 | (0.0-12.1) | -1.7<br>*0.79* | 1.1 | (0.1-2.1) | -3.2<br>*0.43* | 1.0 | (1.0-1.0) | -0.1<br>*0.87* | 0.9 | (0.0-2.1) | -0.1<br>*0.93* | 1.3 | (0.4-2.2) | 0.4<br>*0.64* | 1.5 | (0.0-3.7) | 0.2<br>*0.86* | NA | | . |
| 9. Manufacturing & utilities | 16.9 | (3.9-29.9) | 1.8 | (0.2-3.4) | -15.1<br>*0.02* | 0.5 | (0.0-1.1) | -1.3<br>*0.14* | 6.0 | (0.0-16.6) | 5.5<br>*0.31* | 8.5 | (2.0-15.0) | 2.6<br>*0.66* | 1.3 | (0.2-2.4) | -7.2<br>*0.03* | 4.4 | (0.5-8.2) | 3.1<br>*0.13* | 0.9 | (0.0-1.8) | -3.5<br>*0.08* |

**Pre-COVID:** February 1st 2018 to March 17th 2019**; Spring 2020**: April 21st to May 25th 2020; **Summer 2020**: July 3rd to August 31st 2020; **Fall 2020**: October 1st to December 16th 2020; **Holidays 2020-2021**: December 17th 2020 to January 8th 2021. **1st wave:** Spring 2020; **2nd wave**: September 2020 to February 2021.

***** Mean number of social contacts at work weighted to represent the mean daily contact over one usual week, estimated among workers.

† Diff: Absolute difference with the preceding period and p-value of this difference.

NA: Not available because there was no participant in these types of employment for these specific periods.

**Table S8. Time trends in the proportion of workers who reported working remotely, according to the type of employment (2016 National occupation classification).**

| | Spring 2020<br>n=179 | | Summer 2020<br>n=364 | | | September 2020<br>n=180 | | | Fall 2020<br>n=406 | | | Holidays 2020-21<br>n=395 | | | January 2021<br>n=182 | | | February 2021<br>n=179 | | |
|---|---|---|---|---|---|---|---|---|---|---|---|---|---|---|---|---|---|---|---|---|
| | **%** | **95% CI** | **%** | **95% CI** | **Diff[†]**<br>***p*** | **%** | **95% CI** | **Diff[†]**<br>***p*** | **%** | **95% CI** | **Diff[†]**<br>***p*** | **%** | **95% CI** | **Diff[†]**<br>***p*** | **%** | **95% CI** | **Diff[†]**<br>***p*** | **%** | **95% CI** | **Diff[†]**<br>***p*** |
| All workers* | 54.9 | (47.1-62.7) | 41.7 | (36.0-47.3) | -13.2<br>*0.007* | 44.2 | (36.4-52.0) | 2.6<br>*0.60* | 49.0 | (44.0-54.1) | 4.8<br>*0.31* | 44.1 | (38.8-49.3) | -5.0<br>*0.18* | 41.7 | (34.2-49.3) | -2.3<br>*0.62* | 57.0 | (49.4-64.6) | 15.3<br>*0.005* |
| 0. Management | 73.9 | (55.7-92.0) | 43.1 | (29.2-57.1) | -30.7<br>*0.009* | 55.3 | (35.0-75.6) | 12.1<br>*0.33* | 63.2 | (49.9-76.6) | 8.0<br>*0.52* | 54.9 | (40.1-69.7) | -8.4<br>*0.41* | 30.8 | (12.2-49.4) | -24.1<br>*0.05* | 64.1 | (46.6-81.6) | 33.4<br>*0.01* |
| 1. Business, finance & administration | 70.9 | (54.8-87.0) | 60.7 | (48.1-73.3) | -10.1<br>*0.33* | 63.5 | (47.6-79.5) | 2.8<br>*0.79* | 57.1 | (47.1-67.0) | -6.5<br>*0.50* | 64.6 | (53.4-75.8) | 7.5<br>*0.32* | 77.9 | (62.9-92.9) | 13.3<br>*0.16* | 68.9 | (52.5-85.3) | -8.9<br>*0.43* |
| 2. Natural & applied sciences | 88.3 | (77.1-99.5) | 70.6 | (56.7-84.5) | -17.8<br>*0.05* | 56.7 | (37.1-76.2) | -13.9<br>*0.26* | 68.6 | (56.6-80.5) | 11.9<br>*0.31* | 66.7 | (54.4-79.1) | -1.8<br>*0.84* | 69.4 | (50.7-88.2) | 2.7<br>*0.82* | 77.4 | (64.6-90.1) | 8.0<br>*0.49* |
| 3. Health | 16.5 | (0.0-33.6) | 5.0 | (0.0-10.8) | -11.5<br>*0.21* | 30.1 | (0.0-67.4) | 25.1<br>*0.19* | 14.8 | (2.4-27.2) | -15.2<br>*0.45* | 9.0 | (0.0-18.8) | -5.8<br>*0.47* | 5.4 | (0.0-15.8) | -3.6<br>*0.63* | 16.4 | (0.0-46.0) | 10.9<br>*0.49* |
| 4. Education, law & social, community & gov. services | 69.3 | (44.1-94.5) | 52.0 | (33.8-70.2) | -17.3<br>*0.28* | 48.6 | (25.7-71.4) | -3.4<br>*0.82* | 53.1 | (38.2-68.0) | 4.5<br>*0.75* | 72.3 | (56.5-88.1) | 19.2<br>*0.08* | 68.2 | (47.0-89.5) | -4.1<br>*0.76* | 61.8 | (35.2-88.4) | -6.4<br>*0.71* |
| 5. Art, culture, recreation & sport | 54.9 | (20.6-89.3) | 68.9 | (50.8-87.0) | 14.0<br>*0.48* | 49.1 | (12.7-85.5) | -19.8<br>*0.34* | 78.0 | (60.2-95.8) | 28.9<br>*0.16* | 69.4 | (43.5-95.3) | -8.6<br>*0.59* | 100.0 | (100.0-100.0) | 30.6<br>*0.02* | 89.0 | (67.6-110.4) | -11.0<br>*0.32* |
| 6. Sales & services | 33.5 | (9.6-57.4) | 21.7 | (9.9-33.5) | -11.8<br>*0.39* | 15.8 | (2.2-29.5) | -5.8<br>*0.53* | 27.7 | (14.6-40.8) | 11.8<br>*0.22* | 20.8 | (9.8-31.8) | -6.9<br>*0.43* | 19.2 | (4.7-33.7) | -1.6<br>*0.86* | 31.0 | (11.4-50.6) | 11.8<br>*0.34* |

**Spring 2020**: April 21st to May 25th 2020; **Summer 2020**: July 3rd to August 31st 2020; **Fall 2020**: October 1st to December 16th 2020; **Holidays 2020-2021**: December 17th 2020 to January 8th 2021. **1st wave:** Spring 2020; **2nd wave**: September 2020 to February 2021.

* Very few workers in the domains of trades, transport & equipment operators (#7), Natural resources agriculture & related production (#8), and Manufacturing & utilities (#9) reported working remotely.

[†] Diff: Absolute difference with the preceding period and p-value of this difference.

**Figure S1. Example of the social contact diary.**

### DATE DAY 1

Year ☐☐☐☐ Month ☐☐ Day ☐☐

**List of persons with whom you have been in contact during this first day, from 5 am today to 5 am tomorrow morning.**

| Ini-tials or nick-name | Age (or age group) of the con-tacted person | Sex | | Person's relation with yourself | | | | Place of contact (*check all that apply*) | | | | | | Total duration of contact with the person | | | | | Did you touch his/her skin? | | How often do you have contact with this person in general? | | | | | Ethnicity of this person | | | | | |
|---|---|---|---|---|---|---|---|---|---|---|---|---|---|---|---|---|---|---|---|---|---|---|---|---|---|---|---|---|---|---|---|
| | | Fe-male | Male | House-hold member | Family member not living in house-hold | Friend / Col-league | Other | Home / Car / Private place | Work | Kinder-garden / School / College / Univer-sity | Public trans-port | Lei-sure | Other | less than 5 min | 5 - 14 min | 15 - 59 min | 1h - 4h | more than 4 h | Yes | No | Daily or almost daily | A few times a week | A few times a month | A few times a year or less | First time | White | Black | Asian | His-panic / Latino | Other | Don't know |
| ____ | ☐☐ (-☐☐) | ☐ | ☐ | ☐ | ☐ | ☐ | ☐ | ☐ | ☐ | ☐ | ☐ | ☐ | ☐ | ☐ | ☐ | ☐ | ☐ | ☐ | ☐ | ☐ | ☐ | ☐ | ☐ | ☐ | ☐ | ☐ | ☐ | ☐ | ☐ | ☐ | ☐ |
| ____ | ☐☐ (-☐☐) | ☐ | ☐ | ☐ | ☐ | ☐ | ☐ | ☐ | ☐ | ☐ | ☐ | ☐ | ☐ | ☐ | ☐ | ☐ | ☐ | ☐ | ☐ | ☐ | ☐ | ☐ | ☐ | ☐ | ☐ | ☐ | ☐ | ☐ | ☐ | ☐ | ☐ |
| ____ | ☐☐ (-☐☐) | ☐ | ☐ | ☐ | ☐ | ☐ | ☐ | ☐ | ☐ | ☐ | ☐ | ☐ | ☐ | ☐ | ☐ | ☐ | ☐ | ☐ | ☐ | ☐ | ☐ | ☐ | ☐ | ☐ | ☐ | ☐ | ☐ | ☐ | ☐ | ☐ | ☐ |
| ____ | ☐☐ (-☐☐) | ☐ | ☐ | ☐ | ☐ | ☐ | ☐ | ☐ | ☐ | ☐ | ☐ | ☐ | ☐ | ☐ | ☐ | ☐ | ☐ | ☐ | ☐ | ☐ | ☐ | ☐ | ☐ | ☐ | ☐ | ☐ | ☐ | ☐ | ☐ | ☐ | ☐ |
| ____ | ☐☐ (-☐☐) | ☐ | ☐ | ☐ | ☐ | ☐ | ☐ | ☐ | ☐ | ☐ | ☐ | ☐ | ☐ | ☐ | ☐ | ☐ | ☐ | ☐ | ☐ | ☐ | ☐ | ☐ | ☐ | ☐ | ☐ | ☐ | ☐ | ☐ | ☐ | ☐ | ☐ |
| ____ | ☐☐ (-☐☐) | ☐ | ☐ | ☐ | ☐ | ☐ | ☐ | ☐ | ☐ | ☐ | ☐ | ☐ | ☐ | ☐ | ☐ | ☐ | ☐ | ☐ | ☐ | ☐ | ☐ | ☐ | ☐ | ☐ | ☐ | ☐ | ☐ | ☐ | ☐ | ☐ | ☐ |

**Figure S2. Quebec COVID-19 epidemiology, related physical distancing measures, and CONNECT data periods.**

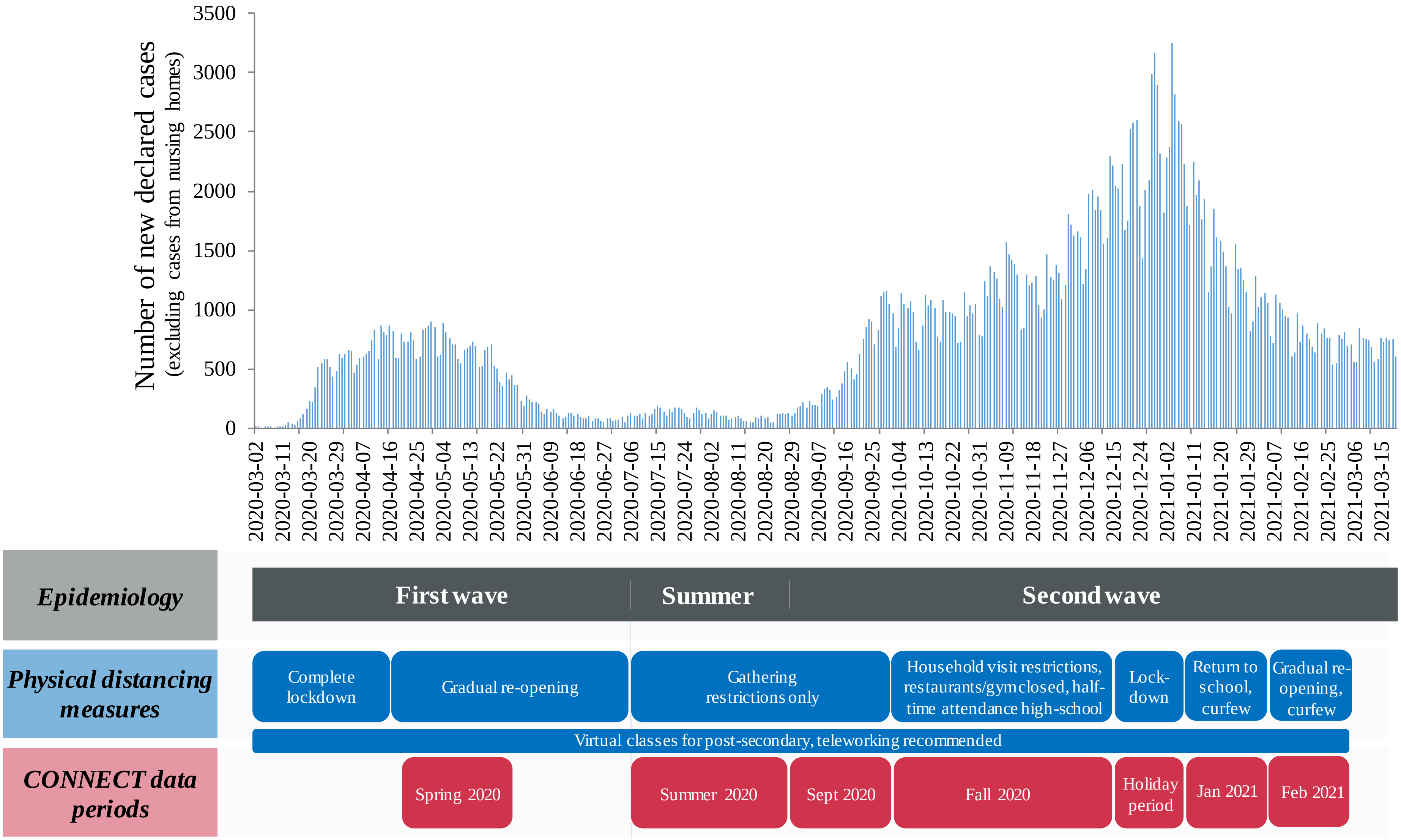

**Figure S3. Flowchart of participant identification for CONNECT 1, CONNECT 2, and CONNECT 3,4,5.**

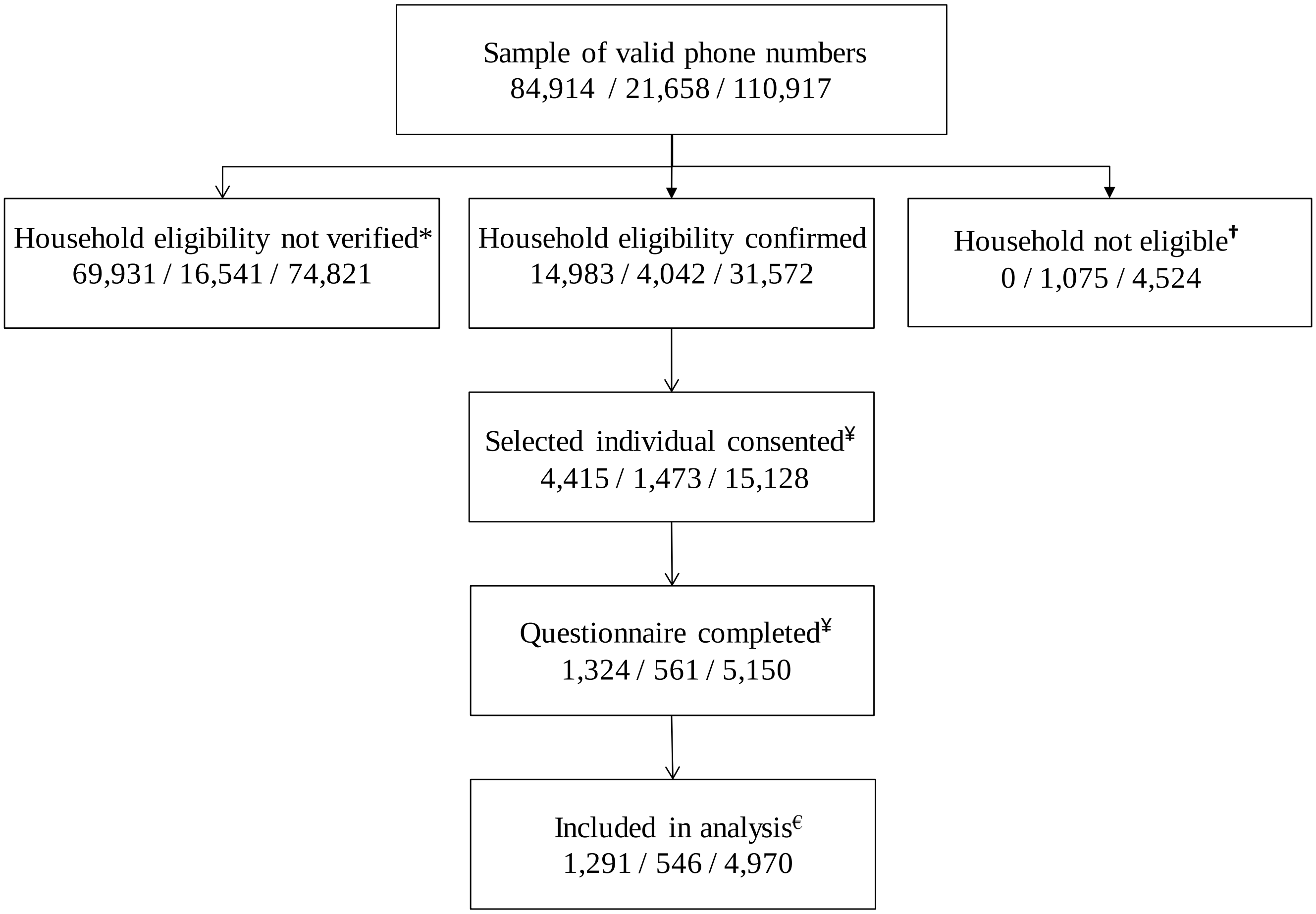

* Household eligibility not verified: no answer, refusal to determine eligibility.

† Household not eligible: out of Quebec.

¥ Participation: questionnaires completed among selected individuals who consented. CONNECT 1: 1,324/4,415 = 30%, CONNECT2: 561/1,473 = 38%, CONNECT3,4,5: 5,150/15,128 = 34%.

€ Participants were excluded from the analysis when the social contact diary was not adequately completed.

**Figure S4. Mean number of social contacts according to the intensity of public health measures in Quebec as summarized by the stringency index.**

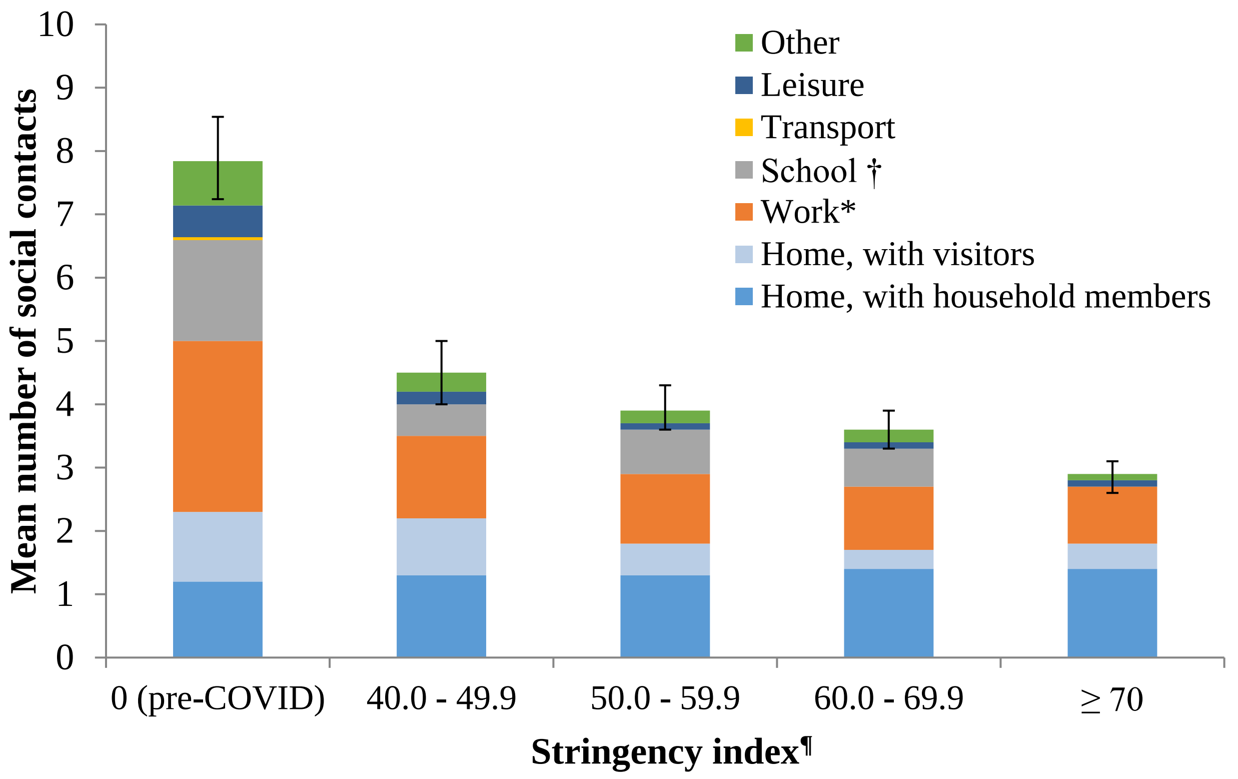

† Contacts for workers in schools were included in the school location.

* Contacts at work were truncated to a maximum of 40 contacts per day.

¶ Stringency index: Higher values indicate stricter measures; the association between social contacts and the stringency index was examined irrespective of periods.

Error bars represent the 95% confidence interval of the total number of social contacts.
